# The link between autism and sex-specific neuroanatomy, and associated cognition and gene expression

**DOI:** 10.1101/2022.03.18.22272409

**Authors:** Dorothea L. Floris, Han Peng, Varun Warrier, Michael V. Lombardo, Charlotte M. Pretzsch, Clara Moreau, Alexandros Tsompanidis, Weikang Gong, Maarten Mennes, Alberto Llera, Daan van Rooij, Marianne Oldehinkel, Natalie Forde, Tony Charman, Julian Tillmann, Tobias Banaschewski, Carolin Moessnang, Sarah Durston, Rosemary J. Holt, Christine Ecker, Flavio Dell’Acqua, Eva Loth, Thomas Bourgeron, Declan G. M. Murphy, Andre F. Marquand, Meng-Chuan Lai, Jan K. Buitelaar, Simon Baron-Cohen, Christian F. Beckmann, the EU-AIMS LEAP Group

## Abstract

**Objectives:** The male preponderance in autism spectrum conditions (ASC) prevalence is among the most pronounced sex ratios across different neurodevelopmental conditions. Here, we aimed to elucidate the relationship between autism and typical sex-differential neuroanatomy, cognition, and related gene expression.

**Methods:** Using a novel deep learning framework trained to predict biological sex, we compared sex prediction model performance across neurotypical and autistic males and females. Multiple large-scale datasets were employed at different stages of the analysis pipeline: a) Pre-training: the UK Biobank sample (>10.000 individuals); b) Transfer learning and validation: the ABIDE datasets (1,412 individuals, 5-56 years of age); c) Test and discovery: the EU-AIMS/AIMS-2-TRIALS LEAP dataset (681 individuals, 6-30 years of age) and d) Specificity: the Neuroimage and ADHD200 datasets (887 individuals, 7-26 years of age).

**Results:** Across both ABIDE and LEAP we showed that features positively predictive of neurotypical males were on average *more* predictive of autistic males (*P*=1.1e-23). Features positively predictive of neurotypical females were on average *less* predictive of autistic females (*P*=1.2e-22). These accuracy differences in autism were not observed in individuals with ADHD. In autistic females the male-shifted neurophenotype was further associated with poorer social sensitivity and emotional face processing while also with associated gene expression patterns of midgestational cell types.

**Conclusions:** Our results demonstrate a shift in both autistic male and female individuals’ neuroanatomy towards male-characteristic patterns associated with typically sex-differential, social cognitive features and related gene expression patterns. Findings hold promise for future research aimed at refining the quest for biological mechanisms underpinning the etiology of autism.

## Introduction

Autism spectrum conditions (henceforth ‘autism’ or ASC) are a set of neurodevelopmental conditions diagnosed four to five times more often in males than in females (1). This male preponderance in prevalence has partially been attributed to an under-recognition of autistic females (2). Despite this, studies accounting for ascertainment bias report a male:female ratio of 3:1 (3) implying the involvement of neurobiological mechanisms in the sex-biased prevalence of autism. This raises the critical question of how quantitative biological factors related to typical sexual differentiation are involved in the neurobiology of autism. Identifying such mechanisms related to biological sex will broaden our understanding of sex-differential etiology of autism.

Studies of biological sex differences in the autistic neural phenotype have gained considerable attention in recent years (4). However, how typical sexual differentiation is associated with the neuroanatomy of autistic males and females remains unclear (5–7). Initial efforts show that autistic females tend to exhibit a brain phenotype resembling that of neurotypical (NT) males (6, 8–11), while findings are more mixed in autistic males (12). It thus remains to be established how a shift towards ‘neurotypical maleness’ differentially informs neurobiological underpinnings in autistic males and females.

Given the neurodevelopmental origins of autism (13), the underlying biological mechanisms are likely to act early ontogenetically. The prenatal period is a critical developmental window in which genetic, hormonal and neuroimmune organizational factors have long-lasting effects on sexual differentiation and pathways implicated in neurodevelopmental conditions such as autism (14). Different etiological models have been put forward explaining the sex-differential autism likelihood (15–17), e.g., the female protective^i^ effect (18–20) and increased steroidogenic activity in prenatal neurodevelopment (15, 16, 21, 22). Thus, a key question is how proposed explanations translate into neurobiological cascades from genome to neural and behavioural phenotype. Specifically, we need to better understand how sex-differential neural phenotypes are linked to cognition and genomic mechanisms, and whether they can be traced back to the prenatal developmental period and specific pathways.

Motivated by the need to address previous shortcomings such as small sample sizes (especially of females) and lack of independent replication, tests of methodological robustness (23) and careful accounting for confounding variables (e.g., brain volume (7)), we aimed to comprehensively establish whether the neuroanatomy in male and female autistic people shows an on-average shift towards (and beyond) the typical male neuroanatomy, and whether this is associated with autism-associated and typically sex-differential cognition and related gene expression. In line with etiological models and evidence suggesting that typical male neurobiology is associated with higher autism likelihood (14, 16, 20, 21), we derived the following hypotheses: using a structural brain imaging-based sex prediction classifier pre-trained in neurotypical males and females to predict male sex, *H1)* within autism, females will be less accurately classified than males; *H2)* within females, autistic females will be less accurately classified than NT females; and *H3)* within males, autistic males will be more accurately classified than NT males. We further examined whether male-shifted neuroanatomy is *a)* specific to autism, b) neurodevelopmentally different across age, *c)* associated with specific brain regions and clinical features, and *d)* linked to gene expression associated with autism, prenatal development, or sex differences.

To test our hypotheses, we employed a novel deep learning framework leveraging several large-scale neuroimaging datasets – the UK Biobank (UKB) (24) comprising over 10,000 individuals, the Autism Brain Imaging Data Exchange (ABIDE (25, 26)) and EU-AIMS/AIMS-2-TRIALS Longitudinal European Autism Project (LEAP (27, 28); the largest European multi-centre initiative aimed at identifying biomarkers in autism) – with proportionally large numbers of autistic females. Based on T1-weighted structural brain images, we first established a sex prediction classifier with high accuracy in UKB, next transferred and validated it in ABIDE, and finally tested it in LEAP to assess sex-classification accuracies in autistic males and females. Thereafter, to establish the specificity of our results to autism, we applied the sex prediction model to an independent sample of individuals with attention-deficit/hyperactivity disorder (ADHD) – another male-biased neurodevelopmental condition.

We next employed a novel framework to translate global prediction accuracies into spatially specific sex predictions at the level of the brain to identify the most implicated brain regions and associated cognitive features. Finally, to understand associated biological processes, we investigated the enrichment of genes highly expressed in regions associated with sex-differential prediction with a range of (prenatal) genes associated with autism or sex. We tested the hypothesis that genes expressed in male-shifted brain regions are enriched for prenatal genes and/or upregulated autism-associated genes related to microglia (29–31) and excitatory neurons (32), and/or differentially expressed by sex or gonadal steroids (33).

## Methods

### Samples

**Pre-training**: We used the preprocessed and quality-controlled first release of UKB (24, 34) comprising 14,503 individuals (7,584 NT females [NT-F], 6,919 NT males [NT-M], 44–80 years) to pre-train the convolutional neural networks (CNN)(35) (see Supplemental Information [SI]). **Validation/transfer**: We selected a sample from the publicly-available ABIDE I (26) and II (36). The selection process (see SI) resulted in a total of 1,412 individuals (115 autistic females [ASC-F], 526 autistic males [ASC-M], 239 NT-F, 532 NT-M, 5–56 years; see ST1; numbers were balanced across sex and diagnosis at all stages of model training, transfer and validation, see SI). **Testing**: We included participants from the LEAP (27, 28) cohort. After selection procedures (see SI), the final sample consisted of 681 individuals (286 ASC-M, 109 ASC-F, 188 NT-M, 98 NT-F, 6–30 years; Table 1 and SF1A; numbers were balanced across sex and diagnosis when testing the model, see SI). For a list of clinical and cognitive measures included in analyses, see SI. **Specificity:** To assess specificity of results, we selected an independent sample of individuals with ADHD based on the publicly-available ADHD200 (37) and the local NeuroImage (38) samples (324 males with ADHD [ADHD-M], 130 females with ADHD [ADHD-F], 225 NT-M, 208 NT-F, 7–26 years; see SI, ST2 and SF1B).

**Table 1.**
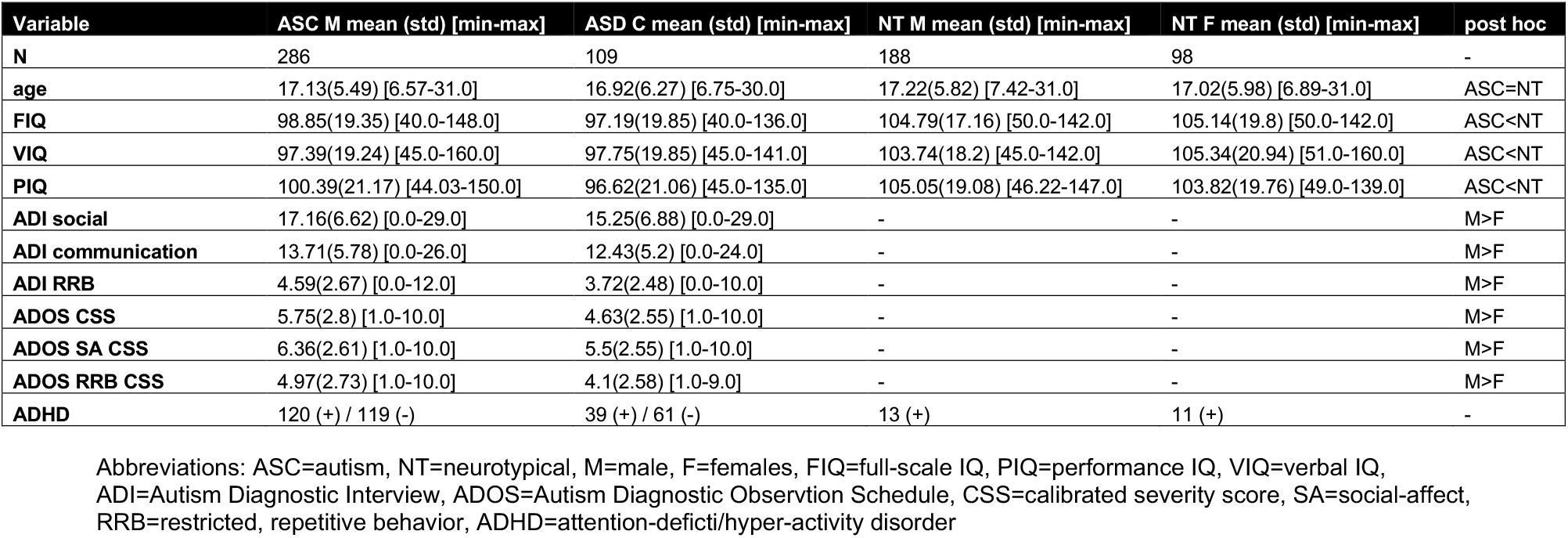
LEAP sample characterization.

### Sex predictions

The detailed steps describing image preprocessing, model training and testing are outlined in the SI and ST3. To summarize, we first pre-trained the CNN model in UKB for sex-classification (i.e., yielding a probability of male sex) using the brain-extracted, bias-corrected and linearly registered (12 degrees-of-freedom) T1-weighted images as input (SF2). Next, we applied the pre-trained models to ABIDE via transfer-learning (39, 40). For this, we randomly re-sampled the ABIDE-cohort 100 times for training and validation using balanced numbers of individuals in each diagnostic (ASC/NT) and sex group (M/F) across all sites (SF1C, SF1E, ST4). We then applied the 100 fine-tuned models to every individual in LEAP – again using sex/diagnostic group-balanced numbers. To summarize prediction results, we computed *a)* individual-level ensemble prediction probabilities for every individual based on the median value of the 100 prediction probabilities which provides an ensemble measure of predictive confidence, and *b)* group-level ensemble prediction proportions (i.e., the true positive rate [correctly classified ASC-M] and the true negative rate [correctly classified ASC-F] which provides an ensemble measure of predictive accuracy, hereafter referred to as ‘sex prediction accuracy’) for each sex/diagnostic group (i.e., ASC-M, ASC-F, NT-M, NT-F). For this, all 100 individual-level prediction probabilities were compared against a sex-classification threshold which was defined by the median prediction probability across NT males and females (since we were interested in sex-specific accuracy differences in autism that differed from the NT baseline reference). This implies that the accuracy of the group-level ensemble sex prediction of female and male individuals was the same within NT (i.e., the true positive and true negative rates were the same). Finally, group-differences in these group-level sex prediction accuracies were computed with one-sampled t-tests and associated Cohen’s *d*.

To identify potential age effects, we further checked the sex prediction accuracies as a function of age. For this, we generated four sliding age bins each spanning 10 years (5-15 years, N=144; 10-20 years, N=204; 15-25 years, N=192; 20-30 years, N=120) and computed the median sex prediction accuracy for each bin and diagnostic/sex group. To test functional relevance of sex prediction accuracies, we investigated the associations with autism-related, clinical and sex-differential cognitive features (SI). The same procedure was also applied to the ADHD sample (SF1D, SF1F and ST4) and to autistic individuals with and without co-occurring ADHD. Finally, we ran several sensitivity analyses to control for model-choice, total brain volume and age (SI).

### Interpreting sex predictions at the brain-level

To interpret which brain regions are most important for the sex-classification, we applied Region-Aligned Prediction (RAP) in LEAP. This novel model interrogation approach predicts labels at the brain region level using feature encodings from an intermediate CNN layer (for details and validation of the RAP-method, see SI). This resulted in spatially specific sex prediction maps where every spatial location received 100 predictions (one per model) for each individual. We used the median value as the ensemble-prediction for each voxel, resulting in one RAP-imaging-map per person. Finally, we used these individual RAP-imaging-maps to compute group-t-maps (ASC-M vs. NT-M; ASC-F vs. NT-F; NT-M vs. NT-F) to discover the spatial group differences related to the sex predictions. We further applied a quartile split on the prediction probabilities in ASC-M and ASC-F to also compare highly misclassified and highly correctly classified individuals on their spatial sex predictions.

### Cognitive decoding

We investigated two specific RAP-imaging-t-maps (ASC-F vs. NT-F and ASC-M vs. NT-M) focussing on regions where ASC-M and ASC-F showed higher male regional predictive probabilities (i.e., *positive* t-values). For this, we used the Neurosynth Image Decoder to visualize the top 100 terms most strongly associated with the two RAP-imaging-t-maps. To identify ROIs with highest male regional predictive probabilities, these maps were cluster-corrected (voxel-level *Z*=3.1, *P*=0.001) and used as masks to extract the average RAP-values for each autistic male/female. These ROI-specific RAP-prediction values were then submitted to general linear models (GLMs) to test the association with different cognitive measures. These were picked post-hoc based on the cognitive domains implicated by the cognitive decoding analyses (see SI). Results were FDR-corrected.

### Gene expression decoding and enrichment analyses

To establish genomic associations, we next examined the gene expression profiles of these RAP-imaging-t-maps (ASC-F vs. NT-F and ASC-M vs. NT-M and NT-F vs. NT-M as reference). To isolate genes that are highly expressed in spatial patterns throughout the brain and similar to the sex prediction RAP-imaging-t-maps, we used the gene expression decoding functionality integrated in Neurosynth (41) and NeuroVault (42). This approach has been previously used in several autism-related studies (32, 33, 43, 44). It uses the six donor brains from the Allen Human Brain Gene Expression atlas (45) and statistically test the similarity between spatial gene expression patterns of all 20,787 protein decoding genes and our input maps. Among the resulting list of genes only those surviving FDR *P*<0.05 and with positive t-statistics (i.e., higher male probability) were retained (SI).

Next, we tested the overlap (i.e., enrichment) of our gene lists with different classes of genes acting prenatally and/or relevant in the context of autism and sexual differentiation. These included *a)* autism-associated genes, including common genetic variants (46), rare genetic variants (47, 48) and transcriptionally dysregulated genes (31, 49, 50); *b)* genes from prenatal cell types (51); and *c)* sex-differentially regulated genes such as genes with sex-differential expression from prenatal brain samples (29, 52) and sex-differentially regulated genes by dihydrotestosterone (DHT) (32, 33, 53) from embryonic neural stem cells, and estrogen (54) (SI). We computed the enrichment odds ratios and associated P-values using hypergeometric tests (33) and only genes surviving FDR *P*<0.05 were considered.

## Results

### Sex prediction accuracies in autism

In ABIDE, at the group-level the proportion of correctly classified ASC-F (72%) was on average 9% lower than that of ASC-M (82%) (***H1***: Cohen’s *d*=1.19, *P*=1.1e-20) and 7% lower than that of NT (79%) (***H2***: *d*=1.25, *P*=3.9e-22). The proportion of correctly classified ASC-M was 3% higher than that of NT (***H3***: *d*=0.48, *P*=1.0e-5) (Figure 1A). These results replicated when testing the 100 ABIDE-fine-tuned models in LEAP (Table 2). Specifically, for the same group comparisons, we found that at the group-level the proportion of correctly classified ASC-F (74%) was on average 12% lower than that of ASC-M (86%) (***H1***: *d*=1.59, *P*=5.5e-29) and 6% lower than that of NT (80%) (***H2***: *d*=1.29, *P*=1.2e-22). The proportion of correctly classified ASC-M was 6% higher than that of NT (***H3***: *d*=1.34, *P*=1.1e-23) (Figure 1B). These results were also reflected in differences in individual-level predictive confidence in LEAP (Figure 1C): after ranking the individual-level ensemble sex prediction probabilities, we compared the top 10% values using a Wilcoxon rank sum test. ASC-F showed lower predictive confidence (75%) than NT (84%) (*W*=2.7, *P*=7.1e-03), while ASC-M showed higher predictive confidence (89%) than NT (84%; *W*=2.1, *P*=3.8e-02). Effect sizes of sex differences in sex prediction accuracies between autistic and NT individuals were largest in younger individuals and sex prediction accuracies improved towards adulthood across all groups (Figure 1D). Finally, ASC-F showed an association between poorer performance on a mentalizing test with typical sex-differential performance (55) (the “Reading the Mind in the Eyes Test”) and higher individual-level male predictive confidence (*t*=-2.36, *P*=0.01, *q*=0.04, see SI and SF3A).

**Figure 1.**
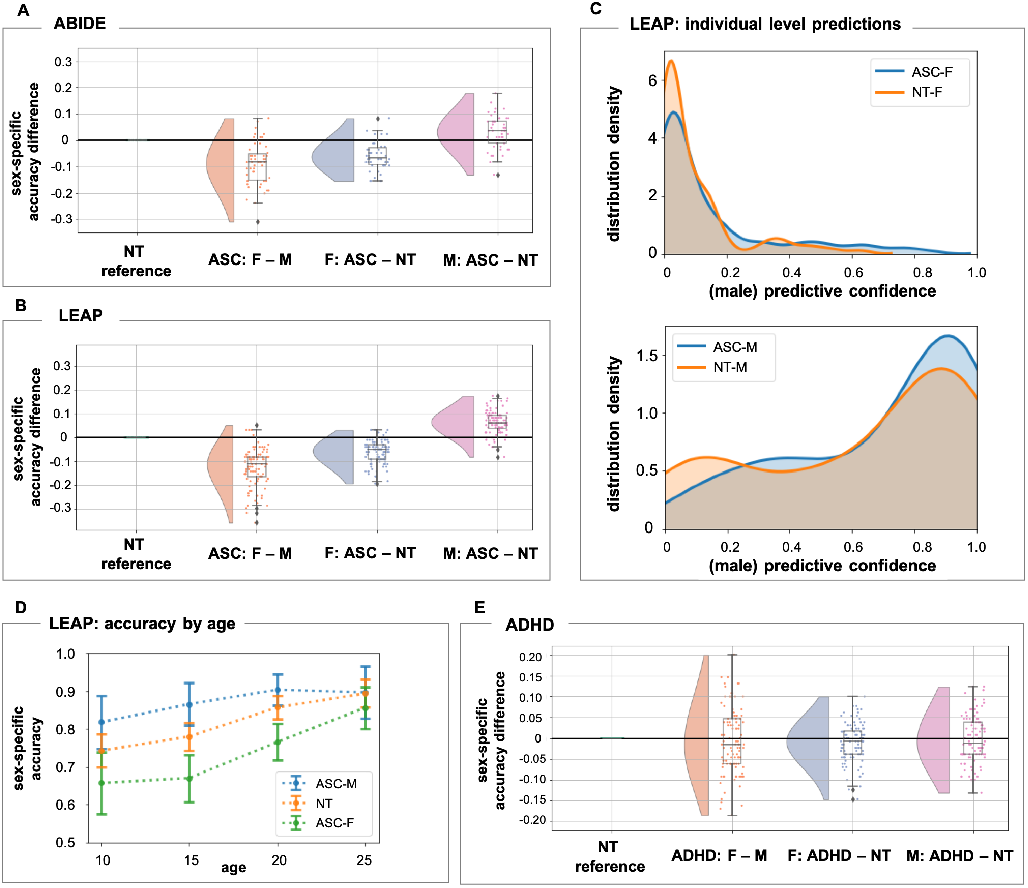
We compared sex prediction accuracy differences between autistic males and autistic females (ASC: F – M), autistic females and neurotypical females (F: ASC – NT) and autistic males and neurotypical males (M: ASC – NT) in both the ABIDE validation sample (**A**) and the LEAP test sample (**B**). Each dot represents one model out of 100 models in total. Negative values mean the model performs worse in the first sex/diagnostic group. **C**) Distribution of individual-level sex prediction accuracies (i.e., predictive confidence of being classified as male) across autistic females and neurotypical females in upper panel and across autistic males and neurotypical males in the lower panel in the LEAP sample. **D**) We generated four sliding age bins each spanning 10 years (5-15 years, N=144; 10-20 years, N=204; 15-25 years, N=192; 20-30 years, N=120) and compared the sex prediction accuracies as a function of increasing age. **E**) Comparison of sex prediction accuracy differences between males and females with ADHD (ADHD: F – M), females with ADHD and neurotypical females (F: ADHD – NT) and males with ADHD and neurotypical males (M: ADHD – NT) in the combined ADHD200 and Neuroimage sample. Abbreviations: NT=neurotypical, ASC=autism, F=females, M=males, ADHD=attention-deficit/hyperactivity disorder.

**Table 2.** sex-specific prediction accuracies across the autism and ADHD samples.

| Cohort |  | ABIDE |  |  | LEAP |  |  | ADHD |  |  |
| --- | --- | --- | --- | --- | --- | --- | --- | --- | --- | --- |
| Group |  | NT | ASC-F | ASC-M | NT | ASC-F | ASD-M | NT | ADHD-F | ADHD-M |
| **Sex-specific accuracy** | Mean | 0.789 | 0.724 | 0.818 | 0.8 | 0.737 | 0.863 | 0.733 | 0.722 | 0.73 |
|  | SD | 0.03 | 0.046 | 0.024 | 0.024 | 0.053 | 0.041 | 0.046 | 0.069 | 0.064 |

Abbreviations: ASC=autism, NT=neurotypical, M=male, F=females, ADHD=attention-deficti/hyper-activity disorder

In the ADHD sample, sex prediction accuracies did not differ across the groups (Figure 1E, Table 2, SI, SF 4 and ST5). Also, in LEAP, the patterns remained the same when stratifying autistic individuals according to co-occurring ADHD (SI, SF5 and ST6). All sets of control analyses confirmed these observed patterns (SI, SF6 and ST7).

### Region-specific sex predictions

The most differentiating regions between NT-M and NT-F were in posterior auditory cortex and frontal-temporal/occipital white matter (WM) pathways (ST8 and Figure 2A). Similarly, the regions with highest shifts towards maleness in ASC-F were also in posterior auditory cortex regions, cerebellum and occipital fusiform gyrus (grey matter [GM]) and corpus callosum and frontal-occipital WM pathways (ST9 and Figure 2B). In ASC-M, regions most similar to NT-M were mostly in subcortical regions (amygdala, pallidum, thalamus), cerebellum, frontal medial cortex and supplementary motor cortex (GM) and in superior fronto-occipital fasciculus (WM) (ST10 and Figure 2C). The NT-M vs. NT-F RAP-imaging-t-map was correlated with the ASC-F vs. NT-F RAP-imaging-t-maps at *r*=0.38, whereas with the ASC-M vs. NT-M RAP-imaging-t-maps at *r*=0.19, further highlighting the similarity between ASC-F and NT-M. For differences across highly mis-classified and correctly classified ASC-F and ASC-M see SF7 and SI.

**Figure 2.**
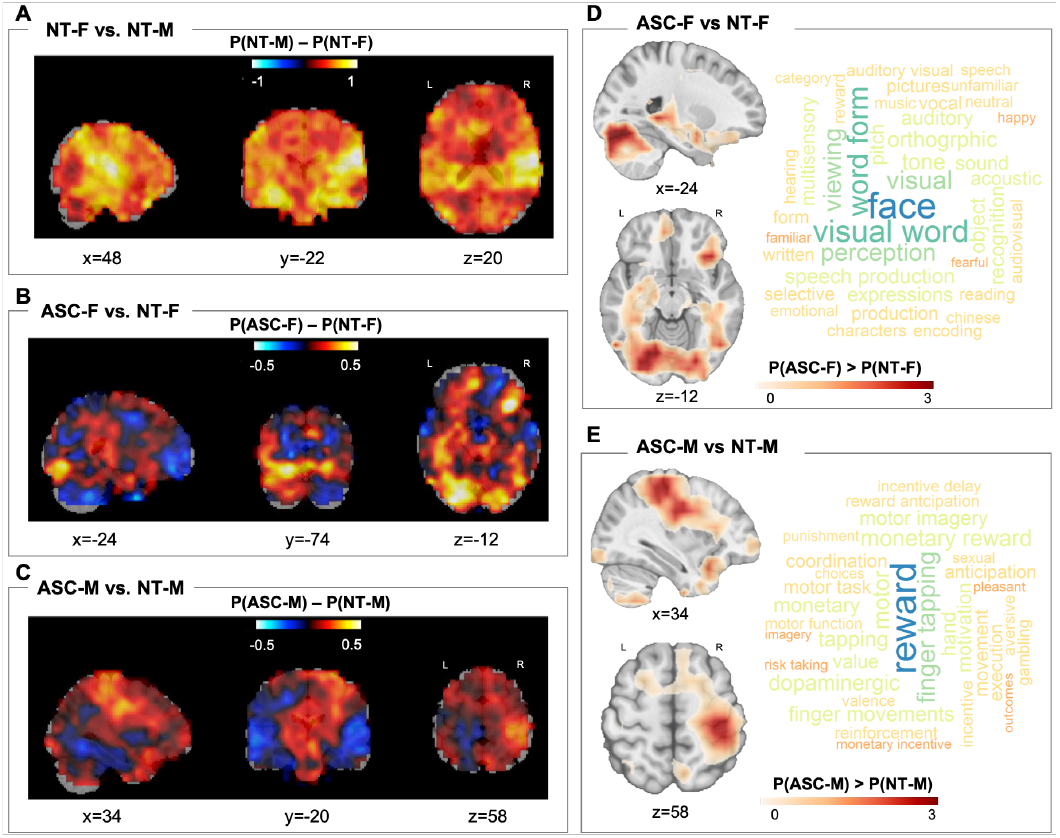
Brain maps A-C depict Cohens-d maps associated with the different RAP-imaging-t-maps (spatial representation of the sex predictions) across **A**) neurotypical females vs. neurotypical males, **B**) autistic females vs. neurotypical females, **C**) autistic males vs. neurotypical males. The colour map encodes the average maleness probability (P; yellow/positive values: higher probability of being classified as male, blue/negative values: higher probability of being classified as female). **D**) Regions where autistic females (ASC-F) showed higher male prediction probabilities than neurotypical females and **E**) where autistic males (ASC-M) showed higher prediction probabilities than neurotypical males (NT-M). To explore the cognitive domains implicated in these (D-E), we used the Neurosynth Image Decoder (http://neurosynth.org/decode/; accessed on January 23^rd^ 2021) to visualize the top 100 terms most strongly associated with the two RAP-imaging-t-maps showing correlations with the imaging maps between r=0.2–0.06. Results showed that the most common cognitive terms associated with the female RAP-t-maps were primarily related to face perception, visual processing and speech in autistic females (ASC-F), whereas to motor and reward processing in autistic males (ASC-M). Abbreviations: NT=neurotypical, ASC=autism, F=females, M=males, P=probability, P(NT-F)=probability of neurotypical females to be classified as males, P(NT-M)=probability of neurotypical males to be classified as males, P(ASC-F)=probability of autistic females to be classified as males, P(ASC-M)=probability of autistic males to be classified as males.

### Cognitive decoding

The most common cognitive terms associated with male-shifted regions in ASC-F were primarily related to face perception, visual processing, and speech, whereas in ASC-M to motor and reward processing (Figure 2D). Based on these results we tested the association between the ROI-specific RAP-based sex prediction values and cognitive measures in LEAP associated with *a)* face processing and communication in ASC-F and *b)* motor and reward processes in ASC-M (see SI for specific measures). While we found no relationships in ASC-M, in ASC-F there was a significant association between predicted maleness and lower accuracy on the Karolinska Directed Emotional Faces task (*t*=-2.6, *P*=0.01, *q*=0.02) (SF3B).

### Gene expression decoding and enrichment analysis

Across both ASC-F and ASC-M, genes that correlated with male-shifted regions showed significant enrichment for a set of transcriptionally *upregulated* genes associated with autism (Figure 3A). In contrast, genes associated with sex-differential regions between NT-M and NT-F showed significant enrichment for transcriptionally *downregulated* autism-associated genes (rightmost panel Figure 3A). Genes correlated with male-shifted regions in ASC-F were significantly enriched for prenatal excitatory neuronal cell types (51) (Figure 3B), and autosomal female differentially-expressed (DE) genes, X-chromosomal male-DE genes and genes downregulated by estrogen (Figure 3C). Strikingly, sex-differential maps in NT-M and NT-F showed similar expression patterns to male-shifted regions in ASC-F (Figure 3A-C middle and rightmost panels). On the other hand, male-shifted regions in ASC-M were significantly enriched for prenatal microglial, progenitor and radial glial cell types (51) (Figure 3B leftmost panel) and upregulated genes by DHT and estrogen (Figure 3C leftmost panel). These results remained largely unchanged when using a more restrictive background total of 16,906 genes (56) (expressed in cortical tissue) (SF8) (44).

**Figure 3.**
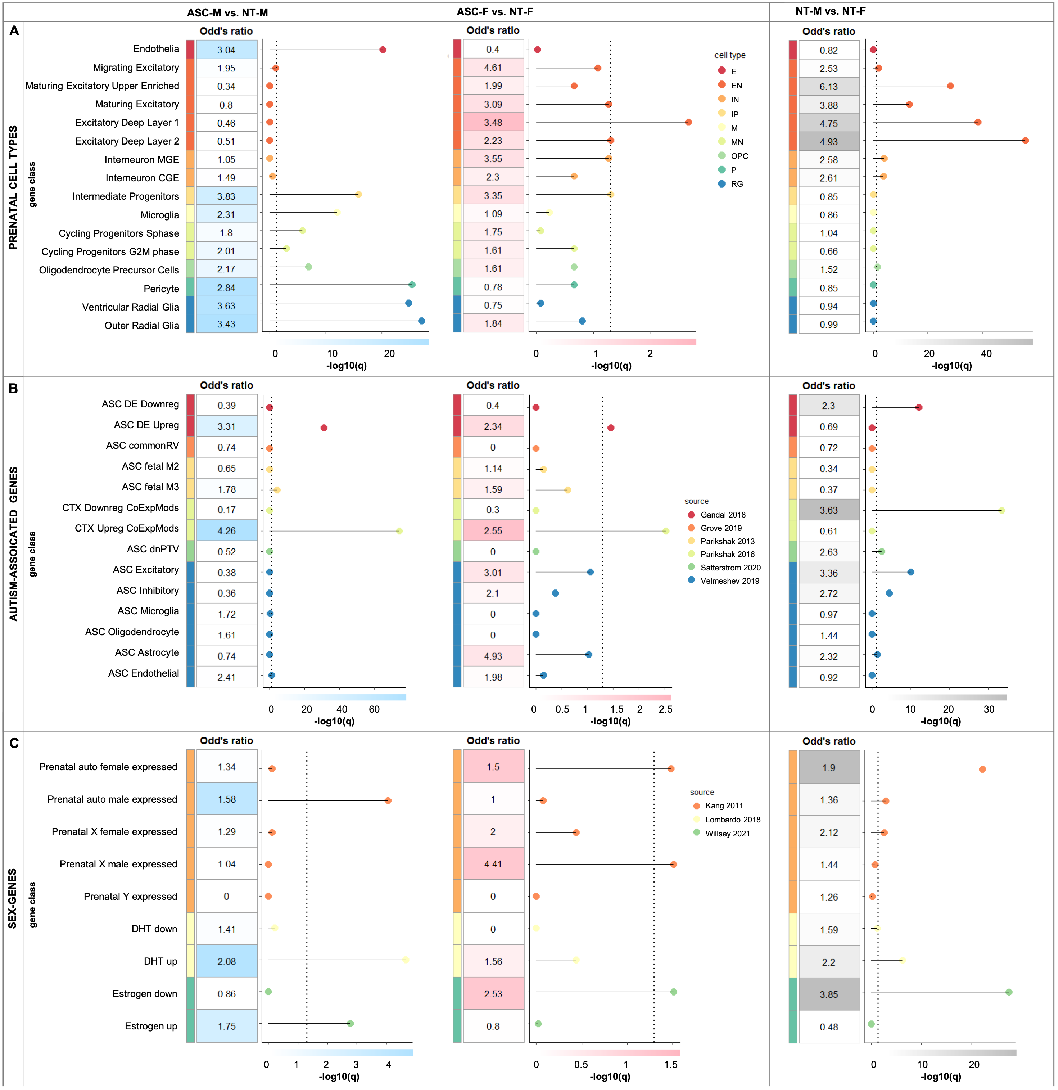
Odds-ratios at an FDR-corrected *P*<0.05 resulting from the gene set enrichment analyses for RAP-imaging-t-maps (ASC-M vs. NT-M; ASC-F vs. NT-F; NT-M vs. NT-F) and associated gene lists with different classes of genes acting prenatally and relevant in the context of autism and sexual differentiation. **A**) autism-associated genes, including common genetic variants (ASC commonRV)(46), de novo mutations (fetal gene co-expression modules [ASC fetal M2, ASC fetal M3](47); 102 rare, de novo protein truncating genes [ASC dnPTV])(48) and transcriptionally dysregulated genes (differentially-expressed downregulated [ASC DE Downreg], differentially-expressed upregulated [ASC DE Upreg](50); cortical downregulated co-expression modules [ASC CTX Downreg CoExpMods], cortical upregulated co-expression modules [ASC CTX Upreg CoExpMods](31); ASC Excitatory, ASC Inhibitory, ASC Microglia, ASC Oligodendrocyte, ASC Astrocyte, ASC Endothelial(49)), **B)** genes from parental cell types(51) (endothelia (E), excitatory neurons (EN; migrating excitatory, maturing excitatory, maturing excitatory upper enriched, excitatory deep layer 1, excitatory deep layer 2), interneurons (IN; interneuron MGE and interneuron CGE), intermediate progenitors (IP), microglia (M), cycling progenitors (MN; cycling progenitor S-phase and cycling progenitors G2M phase), oligodendrocyte precursors (OPC), pericyte (P), and radial glia (RG; ventricular radial glia and outer radial glia), and **C**) sex-differentially expressed genes acting prenatally such as differentially expressed genes by dihydrotestosterone (DHT)(32, 33, 53), estrogen(54) and (autosomal and X-/Y-chromosome-linked) sex-differential gene array expression data from prenatal samples(29, 52). Abbreviations: NT=neurotypical, ASC=autism, F=females, M=males.

## Discussion

Our study demonstrates the overlap of neuroanatomical features characteristic of neurotypical males with those of autistic individuals. This pattern was specific to autism and was not observed in ADHD, pointing to possible different underlying biology in different neurodevelopmental conditions, despite both having a male-predominant prevalence. Overall, autistic females constituted the more critical test case for our hypotheses, likely because masculinizing sex-differentiation effects are less likely to reach a ceiling in females. Consistent with this, we observed an association between greater shifts towards neuroanatomical maleness and cognitive functional difficulties in social sensitivity and emotional face processing in autistic females. Associated genomic expression patterns in ASC-F were highly similar to those differentiating NT-M from NT-F, primarily comprising prenatal cell types (excitatory neurons) and up-regulated autism-associated genes. The findings provide key insights into potential neurobiological and genomic underpinnings associated with male-biased autism prevalence.

We emphasize that the majority of autistic females were *correctly* classified and we cannot derive individual-level predictions of autism given the overlapping distributions across NT and autistic individuals. This highlights the large heterogeneity inherent to autism and the importance of identifying biologically meaningful sub-groups (57, 58). Among these, at least one sub-group is characterized by multivariate features in brain structure that are more similar to those of neurotypical males. These multivariate results of male-shifted whole-brain patterns add to previous mass-univariate neuroimaging studies which identify shifts towards the neurotypical male profile in autistic females across both brain structure (8, 59, 60) and function (11, 23, 61). Such masculinized profile in autistic individuals has previously been reported in specific aspects of cognitive style(62, 63). Our study extends this observation to multivariate, male-shifted characteristics in brain structure. Importantly, the findings are in line with the up-to-date notion that there is no strict sexual dimorphism in human neuroanatomy (64, 65), but brains exhibit a ‘multimorphic’ mosaic of male-/female-like features (66, 67) which can reliably distinguish males from females with above-chance to high accuracy (66, 68, 69). It is important to clarify that autistic individuals do not have an ‘extreme male brain’, but rather that multivariate patterns characteristic of neurotypical males are on average more common in a subset of autistic individuals who are overall shifted towards the male neurophenotype. We previously reported such functional brain mosaic in autistic males who exhibited both shifts towards male- and female-like network-connectivity depending on the neural circuit involved (12).

One striking finding is the similarity of neuroanatomical and genomic patterns between male-shifted regions in ASC-F and regions differentiating NT-M from NT-F. Regions most strongly differentiating NT-M from NT-F overlapped with well-established sex-differential regions in posterior auditory and visual regions and hippocampus (70, 71). These visual, face and language processing areas were also among those that least accurately predicted female sex in ASC-F and have previously been associated with male-shifted patterns in ASC-F (6, 23, 72). Higher predicted maleness was also associated with cognitive difficulties in ASC-F such as poorer mentalizing and emotional face processing, pointing to the clinical relevance of a male-shifted neurophenotype in ASC-F. Furthermore, both sex-differential and the cell-type specific expression patterns in male-shifted regions in ASC-F and in regions differentiating NT-M from NT-F were highly similar to each other. These multi-level findings jointly suggest that cellular mechanisms mediating neurotypical brain sexual differentiation likely play a role in the etiology of autism – especially in females.

Particularly, the influence of prenatal masculinisation via androgens has been suggested to play a mechanistic role in autism etiology (15, 16, 53, 73–75). Other steroid hormones, including estrogens, have also been found to be associated with autism-related outcomes, when cohorts were restricted to males (21, 22, 76). Also, neuroimaging research shows that grey matter volume in posterior auditory regions (exhibiting male-shifts in ASC-F here) and in somatomotor areas (exhibiting male-shifts in ASC-M here) are associated with increased androgen sensitivity (77, 78).

We further observe that gene expressions in male-shifted regions in ASC-M are enriched both for genetic markers of neural progenitor (IP) and radial glial (RG) cell types, as well as genes upregulated by DHT and estrogen. In line with this, it has been shown that DHT increases proliferation of intermediate progenitors and basal radial glia (79). Also, differences in proliferative processes through symmetric cell division in IP and RG lead to atypical cortical expansion of surface area in autism (80–82). These downstream consequence of DHT may impact excitatory neuronal signaling, due to expansion of surface area via increased number of cortical columns and neurons within each column. Furthermore, genes involved in excitatory postsynaptic potentials and autism-associated genes affecting excitatory neuronal lineages are dysregulated by DHT (32, 33), and that subsequently, excitation:inhibition (E:I) imbalance is asymmetrically more affected in autistic males (32).

On the other hand, in ASC-F, male-shifted regions are particularly enriched for later differentiated excitatory neurons and genes downregulated by estrogen, but not for genes upregulated by DHT (as in ASC-M). This is in line with clinical findings indicating that autistic females may be characterised by a relative imbalance between androgens and estrogens, rather than high steroid levels across all pathways (83, 84). Conditions related to androgen/estrogen imbalances in females (e.g., polycystic ovary syndrome) have previously been linked to autism likelihood in both mothers and their children (85–88). Further, estrogen-regulated signalling can influence region-specific neurodevelopmental male-shifts and affect neuronal excitation(89–91), but more studies are needed to demonstrate this in human-derived cell-models. Thus, the enrichment effects in IP, RG and DHT in ASC-M and in excitatory neuronal cell types and estrogen in ASC-F may be relevant to the overall idea that one key emergent phenomenon of brain masculinization is the effect it has on E:I imbalance (92), with potentially different underlying sex-steroid-related mechanisms in autistic males and females (76).

Furthermore, microglial neuro-immunological processes may be major contributors to brain masculinization (14). Werling et al. (29) reported enrichment of male-biased microglial expression with autism-upregulated co-expression modules along with enrichment of female-biased synaptic expression with autism-downregulated co-expression modules, corroborating that molecular downstream pathways regulating neurotypical male development interact with those of autism-associated genes. These observations are consistent with our discoveries that *a)* genes expressed in regions contributing to the male-shift in both ASC-M and ASC-F are enriched for upregulated autism-associated genes mapping onto inflammatory pathways (31) (likely underlying vulnerability mechanisms associated with males (93)), *b)* regions differentiating NT-F from NT-M are enriched for autism-downregulated expression modules (likely underlying protective mechanisms associated with females (93)), *c)* the male-shifted regions in ASC-M are enriched for prenatal microglial cell-types and *d)* downstream transcriptionally dysregulated pathways are rather involved in sex-differential processes than upstream genetic susceptibility mechanisms.

Our findings of reduced and superior sex prediction performance in autistic females and males, respectively, are most pronounced in childhood and decrease throughout development to young adulthood. In the age-bin of 20-30 years, we do not observe classification differences across groups. Similarly, two other studies in samples with mean ages of 22 (7) and 26 (94) years similarly show no sex prediction accuracy differences between autistic and neurotypical adults. We thus attribute observed age-related patterns to the fact that sex-differential and neurodevelopmental processes are dynamically unfolding across the lifespan (65). It remains to be established how experiential, environmental, genetic and organizational and activational hormonal effects interact to differentially contribute to observed patterns across different stages of sexual differentiation (e.g., fetal/neonatal development, puberty and menopause).

Finally, it has been suggested that the male-preponderance in neurodevelopmental conditions may be driven by similar genetic and neuroendocrinological mechanisms. Here, we show that autism is associated with a different sex-differential neuroanatomical profile in males and females than in ADHD. A proposed pathway through which sex-differential prevalence and presentation in ADHD is mediated, may be atypical dopaminergic system function modulated by gonadal hormones (95, 96), such as an increase in the striatal dopamine receptor density in prepubertal development in males, but not females (97). Future research needs to pinpoint which exact sex-related mechanisms shape differential neurophenotypes in different male-biased conditions.

### Strengths and limitations

Our analytical approach has multiple strengths. We employ four of the largest cohorts available for our populations of interest (neurotypical, autism, ADHD), thus ensuring both replicability and specificity of findings. We carefully address potential confounds, confirming that results are not driven by differences in brain volume, age and model choice. Our analyses do not rely on artificial features, but by employing a novel deep CNN model with superior performance(35), we take whole-brain anatomical data as predictive features. Nevertheless, there are several limitations. Even though our samples span wide age-ranges and results imply the likely involvement of neurodevelopmental mechanisms, only when investigating a longitudinal cohort will we be able to make causal inferences. Further, we address neither social, cultural or experiential factors (98) which can influence sex-differential brain development (65), nor gender identity which might differ from sex assigned at birth, especially in autistic individuals (99). In addition, while utilising a highly accurate approach for sex-classification, the high level of non-linearity intrinsic to the CNN model makes straightforward interpretations more difficult compared to simpler linear models. We address this issue by a novel approach for model introspection by means of Region-Aligned Prediction. This approach has the advantage of being able to generate spatially resolved estimates of prediction sex-specific accuracy, but due to the convolutional structure of our model, it achieves this with lower spatial fidelity than the original input data. Yet, being able to spatially resolve the sex prediction accuracy enables the further association analyses with cognition and gene expression.

## Conclusion

Understanding the male-preponderance in autism prevalence requires understanding biological processes involved in brain sexual differentiation (14, 16). Our results suggest that autistic individuals are more likely to show a biologically typical male expression of neuroanatomy. Specifically, in autistic females this shift is associated with typically sex-differential, cognitive features and male-shifted genomic expression patterns. Identifying neuroanatomical, cognitive and genetic inter-relations at the intersection of sex-differential and autism-associated neurobiology will provide promising avenues for research into biological mechanisms underpinning autism etiology. Echoing recent calls for an increased inclusion of females in basic neuroscience research (100), we extend this to clinical neuroscience where increased attention to females in neurodevelopmental research will help elucidate sex-differential likelihood factors, as well as potential sex-specific diagnosis and support that are informed by neurobiology.

## Data Availability

Reproducible code can be found at https://github.com/ha-ha-ha-han/UKBiobank_deep_pretrain.

http://fcon_1000.projects.nitrc.org/indi/abide/abide_II.html

https://www.ukbiobank.ac.uk

https://www.aims-2-trials.eu

http://fcon_1000.projects.nitrc.org/indi/adhd200/

## Acknowledgements

We thank all participants and their families for participating in the studies that contribute to the datasets used in this research. We also gratefully acknowledge the contributions of all members of the EU-AIMS/AIMS-2-TRIALS LEAP group: Jumana Ahmad, Sara Ambrosino, Bonnie Auyeung, Sarah Baumeister, Sven Bölte, Carsten Bours, Michael Brammer, Daniel Brandeis, Claudia Brogna, Yvette de Bruijn, Bhismadev Chakrabarti, Ineke Cornelissen, Daisy Crawley, Guillaume Dumas, Jessica Faulkner, Vincent Frouin, Pilar Garcés, David Goyard, Lindsay Ham, Hannah Hayward, Joerg Hipp, Mark H. Johnson, Emily J.H. Jones, Xavier Liogier D’ardhuy, David J. Lythgoe, René Mandl, Luke Mason, Andreas Meyer-Lindenberg, Nico Mueller, Bethany Oakley, Laurence O’Dwyer, Bob Oranje, Gahan Pandina, Antonio M. Persico, Barbara Ruggeri, Amber Ruigrok, Jessica Sabet, Roberto Sacco, Antonia San José Cáceres, Emily Simonoff, Will Spooren, Roberto Toro, Heike Tost, Jack Waldman, Steve C.R. Williams, Caroline Wooldridge, and Marcel P. Zwiers. This project has received funding from the Innovative Medicines Initiative 2 Joint Undertaking under grant agreement No 115300 (for EU-AIMS) and No 777394 (for AIMS-2-TRIALS). This Joint Undertaking receives support from the European Union’s Horizon 2020 research and innovation programme and EFPIA and AUTISM SPEAKS, Autistica, SFARI. Any views expressed are those of the author(s) and not necessarily those of the funders. This work was also supported by the Netherlands Organization for Scientific Research through Vidi grants (Grant No. 864.12.003 [to CFB] and Grant No. 016.156.415 [to AFM]); from the FP7 (Grant Nos. 602805) (AGGRESSOTYPE) (to JKB), 603016 (MATRICS), and 278948 (TACTICS); and from the European Community’s Horizon 2020 Programme (H2020/2014-2020) (Grant Nos. 643051 [MiND] and 642996 (BRAINVIEW). This work received funding from the Wellcome Trust UK Strategic Award (Award No. 098369/Z/12/Z) and from the National Institute for Health Research Maudsley Biomedical Research Centre (to DM). DLF is supported by funding from the European Union’s Horizon 2020 research and innovation programme under the Marie Skłodowska-Curie grant agreement No 101025785. M-CL is supported by a Canadian Institutes of Health Research Sex and Gender Science Chair (GSB 171373), an Academic Scholars Award from the Department of Psychiatry, University of Toronto, and the Centre for Addiction and Mental Health Foundation. MVL is supported by funding from the European Research Council (ERC) under the European Union’s Horizon 2020 research and innovation programme under grant agreement No 755816 (ERC Starting Grant - AUTISMS). SB-C and VW were funded by the Autism Research Trust, the Wellcome Trust, the Templeton World Charitable Foundation. SB-C was also funded by the NIHR Biomedical Research Centre in Cambridge, during the period of this work. We thank Dr. Nicolas Langer and Dr. Amber Ruigrok for helpful feedback on the manuscript.

## Footnotes

i We use the term ‘female protective effect’ as it has been used in the previous literature but note that we do not think it is necessarily appropriate to talk about autism in terms of ‘risk’ or ‘protective’ factors as autism is both a disability and a difference. We suggest replacing the term ‘female protective effect’ with ‘factors reducing likelihood in females’ and replacing ‘risk factors’ with ‘likelihood factors’.

