## Supplemental Material for "The link between autism and sex-specific neuroanatomy, and associated cognition and gene expression"

### **Supplementary Information**

### UK Biobank

#### Participants, study design and exclusion criteria

UK Biobank (UKB) is a large-scale, population-based, biomedical, epidemiological study comprising around 500,000 predominantly neurotypical (NT) participants. A subset among these has contributed brain imaging data (1). Three dedicated imaging centres are equipped with identical scanner models (3T Siemens Skyra, software VD13) for brain imaging scanning using the standard Siemens 32-channel receive head coil. For details on the brain MRI protocols, see Table 1 in Alfaro-Almagro et al., 2018 (2). In this study, we used the T1-weigthed anatomical MR data from 14,503 individuals (mean age 52.7 years, standard deviation 7.5 years, range 44-80 years), of which 12,949 were used for training, 518 for validation and 1,036 for testing. The image preprocessing pipeline and automated quality control steps are described in detail in (2). We used data as preprocessed already (by our laboratory on behalf of UK Biobank), and as available to all researchers who have been granted access to UKB data. The input data to the deep neural network model was brain-extracted, bias-corrected and registered to MNI152 standard space using affine registration as implemented by FSL-FIRST (3).

### ABIDE

#### Participants, study design and exclusion criteria

We combined the Autism Brain Imaging Data Exchange (ABIDE) repositories I (4) (<http://fcon_1000.projects.nitrc.org/indi/abide/abide_I.html>) and II (5) (<http://fcon_1000.projects.nitrc.org/indi/abide/abide_II.html>) for analysis. Sites with same/highly similar imaging acquisition protocols (as outlined and recommended on the website) were merged across ABIDE I and ABIDE II into one single site (KKI and ABIDEII-KKI; NYU and ABIDEII-NYU; SDSU and ABIDEII-SDSU; UCLA_1 and ABIDEII-UCLA_1). Sites providing less than four individuals (referred to as an individual’s imaging data) per sex/diagnostic group were excluded (namely, ABIDEII-BNI_1, ABIDEII-ETH_1, ABIDEII-OILH_2, ABIDEII-SU_2, ABIDEII-TCD_1, ABIDEII-USM_1, ABIDEII-U_MIA_1, CMU, LEUVEN_1, LEUVEN_2, MAX_MUN, OHSU, OLIN, SBL, TRINITY, UCLA_2, UM_2, and USM). Detailed information on imaging acquisition parameters can be found on the above website. Further, we excluded participants with (clinically non-significant) brain atypicalities (N=2), excessive head motion (N=99) and corrupted image quality (N=3).

For the autism datasets, autism diagnosis was determined by clinician’s consensus supported by either one or both ‘gold-standard’ diagnostic instruments, i.e., an Autism Diagnostic Observation Schedule (ADOS) (6) and/or the Autism Diagnostic Interview-Revised, (ADI-R) (7) in all sites but two (UCD and Stanford sites only used diagnostic cut-offs of ADOS and/or ADI-R for inclusion). This selection process resulted in a total of 1,412 individuals including 115 autistic females, 526 autistic males, 239 NT females, and 532 NT males between 5 and 56 years of age across 15 sites. Individuals were matched for age across and for FIQ within diagnostic groups. For further details, see ST2. For details on demographics, see ST1.

### EU-AIMS LEAP

#### Participants, study design and exclusion criteria

All participants with autism had an existing clinical diagnosis of autism according to DSM-IV (8), DSM-IV-TR (9), DSM-5 (10) or ICD-10 (11) criteria. Participants underwent comprehensive clinical, cognitive and MRI assessment at one of six collaborating sites: the Institute of Psychiatry, Psychology and [Neuroscience](https://www.sciencedirect.com/topics/neuroscience/neurosciences), King’s College London (KCL), London, United Kingdom; Autism Research Centre at the University of Cambridge, Cambridge, United Kingdom; Radboud University Nijmegen Medical Centre, Nijmegen, the Netherlands; University Medical Centre Utrecht, Utrecht, the Netherlands; Central Institute of Mental Health, Mannheim, Germany; and University Campus Bio-Medico, Rome, Italy. Exclusion criteria included the presence of any MRI contraindications (e.g., metal implants, braces, claustrophobia) or failure to give informed written consent to MRI scanning, as well as significant hearing or visual impairments not corrected by glasses or hearing aids. In addition, we excluded participants with missing T1-weighted MRI scans, clinically non-significant brain atypicalities (N= 21), and excessive head motion (N=29). The study was approved by the local ethical committees of participating centers, and written informed consent was obtained from all participants or their legal guardians (for participants<18 years). The final sample comprised 395 autistic individuals (286 males and 109 females), and 297 neurotypical controls (188 males and 98 females) between 6 and 30 years of age. Individuals were matched for age across and for FIQ within diagnostic groups. For details see Table 1 and SF1A.

#### Clinical, cognitive and demographic measures

##### Intellectual functioning

General intellectual abilities were assessed using the **Wechsler Abbreviated Scales of Intelligence**-Second Edition (WASI-II (12)), or if unavailable, the Wechsler Intelligence Scale for Children-III/IV (WISC-III/IV (13, 14)) for children or Wechsler Adult Intelligence Scale for Adults-III/IV (WAIS-III/IV (15, 16)) for adults. Standardized estimates of verbal IQ (VIQ), performance IQ (PIQ), and full-scale IQ (FIQ) were derived using IQ norms with mean=100 and SD=15.

##### Autism-associated, clinical features

The **Autism Diagnostic Observation Schedule** (ADOS-G (6)) was used to measure current, clinically observed core symptoms of autism. Based on the updated algorithm totals (17, 18), we report Calibrated Severity Score (CSS) for ‘Social Affect’ indexing social-communication difficulties and ‘RRBs’ indexing restricted and repetitive behaviours. CSS Total serves an overall indicator of autism severity. The CSS ranges from 1 to 10, with higher scores indicating more substantial autism symptom severity. For follow-up analyses (see relative importance section) we median-split the CSS measure into a group with high CSS and one with low CSS.

The **Autism Diagnostic Interview-Revised** (ADI-R (7)) is a semi-structured caregiver interview completed by parents or caregivers. Algorithm scores were derived from current and historical symptom information for the domains of Reciprocal Social Interaction, Communication, and Restricted, Repetitive and Stereotyped Behaviours and Interests.

**The Social Responsiveness Scale**, Second Edition (SRS‐2 (19)) is a quantitative measure of autistic traits and is composed of 65 items asking about characteristic autistic behaviour over the previous six months. The total raw score is transformed into sex-specific T scores. Parent report was used for all autistic participants and autistic adults additionally completed the self-report form.

The **Repetitive Behaviour Scale**‐Revised (RBS‐R (20)), composed of 43 items, was used to derive parent‐reported total raw scores for restricted and repetitive behaviours, with higher scores indicating a greater level of atypical behaviours.

Sensory processing atypicalities were assessed using the **Short Sensory Profile** (SSP (21)) across 38 items, from which a total raw score was obtained (lower scores indicate more atypicality) that reflect dysfunction across multiple sensory domains.

Adaptive behaviour was assessed with semi-structured parent/caregiver interviews using the **Vineland Adaptive Behaviour Scale**-Second Edition (22). This measures a person’s current level of functioning across three domains: communication (expressive, receptive, and written), daily living skills (community, domestic, and personal), and socialization (coping skills, interpersonal relationships, and play and leisure time). For each domain, standard scores were obtained and combined to generate an Adaptive Behavior Composite (ABC) score. Standard scores have a mean of 100 (SD=15), with lower scores indicating greater functional impairment.

##### Co-occurring conditions (ADHD)

**Attention-deficit/hyperactivity disorder** (ADHD) symptoms were assessed with the DSM-5 ADHD rating scale, covering both inattention and hyper-activity/impulsivity symptoms based on either self- or parent-report (23). Self-report scores were only used when parent-report scores were unavailable (N=83). A categorical variable was computed based on the DSM-5 criteria (i.e., at least five positive responses in children and six in adults on either or both scales).

##### Sex-differential cognitive measures

The **Autism Spectrum Quotient** (AQ) (24) is a self- or parent-reported questionnaire that aims to quantify autistic traits. Depending on their age (adults 18-30, adolescents 12-17, children 6-11) and ability level, participants received either an adult (50 items), adolescent (50 items) or child (50 items) version of the test. Before merging the three different versions across all individuals, each version was z-standardized within each diagnostic group separately.

The **Empathy Quotient** (EQ) was used to measure a cognitive style described as ‘the drive to identify a person's thoughts and feelings and to respond to these with an appropriate emotion’. The **Systemizing Quotient** (SQ) was used to measure a cognitive style characterised by the motivation to predict lawful events (using if-then rules) and observations of input-operation-output relationships and includes good attention to detail. For both, age-appropriate versions were used for children (25), adolescents (26) and adults (27, 28). Before merging the three different versions across all individuals, each version was z-standardized within each diagnostic group separately.

The **Reading the Mind in the Eyes** test (29) asks participants to identify complex emotions and mental states based only on the eye region of a face. Depending on their age (adults 18-30, adolescents 12-17, children 6-11) and ability level, participants received either an adult (36 items), adolescent (31 items) or child (28 items) version of the test. Percentage of correct answers was used as the outcome variable. Before merging the three different versions across all individuals, each version was Z-standardized within each diagnostic group separately.

##### Other cognitive measures (selected post-hoc)

**Social and monetary reward task**

Participants performed a social and monetary incentive delay task within the MRI scanner. Participants were asked to give a speeded response (button press) to a visual target screen flash. For all detail on this task, refer to (30). Behavioural performance on the two tasks was extracted as separate accuracies (i.e., percentage of successful trials) and used as independent variables for our analyses.

**Emotional face matching task (Hariri)**

Participants completed a well-established face matching task (31) within the MRI scanner, with alternating blocks of faces (showing angry and fearful emotions) and control conditions. In the emotional face condition, a target face has to be matched to one of two probes (identity match) by pressing the left or right button of a response device. Analogously, in the control condition, participants are asked to match a target shape (circle or ellipses) to two test shapes. Behavioural performance on the task was extracted as the accuracy on performing the task (i.e., percentage of successful trials) and used as independent variable for our analyses.

**Karolinska Directed Emotional Faces task**

The Karolinska Directed Emotional Faces (KDEF) task (32, 33) tests for recognition of basic emotions. Participants were administered an adapted version with 70 trials(34) reduced from the original 140 trials, to reduce assessment time. In each trial, participants were shown a photograph of a person’s face and asked to select which of seven words (happy, sad, angry, surprised, afraid, disgusted, or neutral) best describes the expression displayed. Behavioural performance was defined as the accuracy on the task.

#### MRI data acquisition

MRI data were acquired on 3T scanners: General Electric MR750 (GE Medical Systems, Milwaukee, WI, USA) at Institute of Psychiatry, Psychology and Neuroscience, King’s College London, United Kingdom (KCL); Siemens Magnetom Skyra (Siemens, Erlangen, Germany) at Radboud University Nijmegen Medical Centre, the Netherlands (RUNMC); Siemens Magnetom Verio (Siemens, Erlangen, Germany) at Autism Research Centre at the University of Cambridge, United Kingdom (UCAM); Philips 3T Achieva (Philips Healthcare Systems, Best, The Netherlands) at University Medical Centre Utrecht, the Netherlands (UMCU); GE Medical Systems Signa HDxTt at the Rome University; and Siemens Magnetom Trio (Siemens, Erlangen, Germany) at Central Institute of Mental Health, Mannheim, Germany (CIMH). Procedures were undertaken to optimize the MRI sequences for the best scanner-specific options, and phantoms and travelling heads were employed to assure standardization and quality assurance of the multi-site image-acquisition. Structural images were obtained using a 5.5-minute MPRAGE sequence (TR=2300ms, TE=2.93ms, T1=900ms, voxels size=1.1x1.1x1.2mm, flip angle=9°, matrix size=256x256, FOV=270mm, 176 slices). For further details see ST3.

### Preprocessing

For imaging and data acquisition parameters in UKB see (2), in ABIDE see http://fcon_1000.projects.nitrc.org/indi/abide/ and in LEAP see ST3. The UKB MRI data have been preprocessed using a standard processing pipeline specified in (2). We used the brain-extracted, bias-corrected and linearly registered (12 degree-of-freedom; to MNI152 standard space) input images to train the convolutional neural network (CNN) / Simple Fully Convolutional Network (SFCN). For ABIDE, LEAP and the ADHD samples, structural T1-weighted images were preprocessed using a standard preprocessing pipeline that included tools from the FMRIB Software Library (FSL version 5.0.6 and 6.0; <http://www.fmrib.ox.ac.uk/fsl>). After brain extraction using FSL BET (35) and bias-correction, T1-weighted images of each participant were linearly registered to MNI152 standard space using 12-parameter affine transformation. Importantly, brain volumes of all individuals were normalized in this linear registration step.

### Sex-Classification

#### Simple Fully Convolutional Network (SFCN)

We used a previously validated (36) convolutional neural network (CNN) architecture to estimate biological sex based on the preprocessed imaging data. This architecture is based on VGGNet(37) using a fully convolutional structure (38). As previously reported (36) we keep the number of layers to a minimum which significantly reduces the number of parameters to 3 million and consequently also the computational complexity and memory cost. This lightweight deep learning model is referred to as Simple Fully Convolutional Network (SFCN).

The model consists of seven blocks, as shown in 2. Each of the first five blocks contains a 3-by-3-by-3 3D convolutional layer, a batch normalisation layer(39), a max pooling layer and a ReLU activation layer (40). The 1mm-input-resolution 160 × 192 × 160 3D input image (with little or no brain tissue loss) goes through each block sequentially, with its feature map generated and spatial dimension reduced to 5 × 6 × 5 after the fifth block. The sixth block contains a 1 × 1 × 1 3D convolutional layer, a batch normalisation layer and a ReLU activation layer. The seventh block contains an average pooling layer, a dropout layer (only used for training, with 50% random dropout rate) (41), a fully connected layer and a softmax output layer. The channel numbers used in each convolution layer are [32, 64, 64, 64, 64, 64, 2]. Reproducible code can be found at <https://github.com/ha-ha-ha-han/UKBiobank_deep_pretrain>.

#### Pre-training in UKB

We pre-trained four CNN models in UKB for sex-classification using the brain-extracted, bias-corrected and linearly registered T1-weighted images as input (SF2). The use of such whole brain anatomical data instead of artificial features has been shown in our previous study to have comparable predictive performance across different modalities when using our deep learning model (see Table 5 in (36)). Next, the 14,503 individual brains used for training and validation were randomly divided into four folds for cross-validation, resulting in four CNN models. Each fold contained about one fourth of individuals for testing, and 518 individuals for validation, with remaining individuals used for training. For detailed description of this previously applied training protocol, see (36). All analyses were implemented using Python and PyTorch(42). Final results revealed a sex prediction accuracy of p=99.5% in both males and females in UKB (36).

#### Transfer-learning and validation in ABIDE

Transfer-learning is widely used in medical image analysis given its utility in real world settings where it is expensive or impossible to recollect training data. It does not require the training and test data to be in the same feature space and to have the same distributions. For 2D images, it is common to transfer the models trained with natural images (ImageNet) to medical image datasets (43, 44) to improve the model performance. Similarly, for the 3D MR images, we can use the model pre-trained in a large cohort, and then transfer to smaller samples with different population distribution and different scanner setup, while retaining good predictive performance. Here, we applied transfer learning (45, 46) to transfer the deep learning models trained in UKB to NT individuals in ABIDE. Thus, here, we used the model pre-trained in a large cohort (UKB) and transferred it to a smaller sample (ABIDE) with different distributions (such as age and site) while retaining good predictive performance (44, 46).

##### Balanced samples across sex, diagnosis and site

To use the data in the most unbiased way for sex-classification in ABIDE (and LEAP/ the ADHD sample further on), we randomly sampled the cohort 100 times for training and validation using the following protocol to get balanced numbers of individuals in each diagnostic (ASC/NT) and sex group (M/F) across all sites: (1) sex matching within each site: we sampled the same number of males and females within NTs, and the same number of autistic males and females; (2) selecting training and validation set within each sex group: If the selected number (N) of NTs was more than twice the number of autistic individuals, then we used all of the autistic individuals for validation (N ASC), and the same number of NTs for testing (N ASC), while the remaining N of NTs for training (N NT – N ASC). If the selected number of NTs was less than twice the number of autistic individuals, then we used half of the number of NTs for training (N ASC), the remaining half of NTs for validation (N NT – N ASC), and the same number of autistic individuals for validation (see ST4).

##### Sex predictions in ABIDE

After this individual sampling, the SFCN models were initialized with the UKB-pre-trained weights (each initialization is shared by 25 models), and next, stochastic gradient descend was used to optimize a loss function of cross-entropy for sex-classification. The learning rate was initially 1e-4 and then decayed by a factor of 10 for every 15 epochs (during one epoch, every training individual is seen once by the model), and the final model was obtained after 50 epochs. The trained model was then used to predict sex for the validation set. For each of the 100 models, the sex-classification accuracy was computed for each sex and diagnostic group (i.e., ASC-M, ASC-F, NT-M, NT-F; for details see below).

##### Comparing sex prediction accuracies across diagnostic/sex groups

Since we were interested in sex-specific accuracy differences in autism that differ from the reference (NTs), we took the median sex prediction probability (i.e., predictive confidence) of the NT individuals as the sex-classification threshold (i.e., the median of the balanced neurotypical sample as baseline reference). Specifically, this means that all 100 individual-level predictions were compared against a sex-classification threshold defined by the median prediction probability across NT males and females. An individual-level prediction probability value greater than the median of NT males and females was classified as ‘male’ vs. a value smaller than the median of the NT males and females was classified as ‘female’. This result was than compared against the real biological sex of each individual and summarized in a Boolean value (True=1 or False=0). The true positive rate (i.e., correctly classified ASC-M) was computed by adding up the true positive values and dividing them by true positive + false negative values, while the true negative rate (i.e., correctly classified ASC-F) was computed by adding up the true negative values and dividing them by true negative + false positive values. Resulting sensitivity and specificity are referred to as sex prediction accuracies within each diagnostic/sex group (ASC-M, ASC-F and NTs). This also implies that the sex prediction accuracy of female and male individuals (used as reference) was the same within the NT cohort. In other words, the true positive rate (in NT-M) and the true negative rate (in NT-F) was the same as the balanced accuracy in NT. Finally, in order to compute differences in the group-level sex prediction accuracies across diagnostic/sex groups, we compared the sex prediction accuracy within each balanced (described above) diagnostic and sex group (i.e., ASC-F vs. ASC-M, ASC-F vs. NT(-F) and ASC-M vs. NT(-M)) with one-sampled t-tests and associated Cohen’s *d*.

#### Sex predictions in LEAP and ADHD

After getting the 100 fine-tuned models from ABIDE, we applied the models to every individual in LEAP. The median value of the 100 predictions for every individual was used to form a final individual-level ensemble-prediction. Again, in order to use the balanced neurotypical sample as baseline reference, the prediction threshold was set to the median prediction probability derived from NT males and NT females. We thus compared the sex prediction accuracy within each diagnostic and sex balanced group, namely, ASC-F vs. ASC-M, ASC-F vs. NT(-F) and ASC-M vs. NT(-M).

The same procedure was next applied to the ADHD sample (see SF1D, SF1F and ST4). In order to ensure comparability, we also computed classification accuracies across autistic individuals (from LEAP) with and without a co-occurring diagnosis of ADHD.

#### Region-Aligned Prediction (RAP)

Usually, deep learning models can be considered black boxes that do not provide any information on which brain features are most important for classification. Thus, to identify the most predictive and biologically meaningful features that drive our prediction at the level of the brain, here we employ a novel model interrogation approach – so called Region-Aligned Prediction (RAP). This method generates spatially resolved estimates of sex prediction accuracy and predicts labels at the brain region level using feature maps from an intermediate CNN layer based on the pre-aligned input data. The RAP method aligns the intermediate feature maps (mp4 in this case, corresponding to the 4th layer of the SFCN) of all individuals in the dataset (including both the training and validation sets), and extracts the feature matrix at one spatial location at a time for all individuals. Next, a logistic regression model is trained using the training set feature matrix and applied to the validation set. More specifically, after a CNN model is trained in the typical fashion, we extract feature encodings for one spatial location of all individuals (note, that we do not refer to voxels but spatial locations here, because the input space becomes increasingly abstract across layers and RAP uses features encodings from the intermediate layer). These feature encodings are then used in simple L2-regularized multiple regression to predict sex. This process is repeated independently for every spatial location of the intermediate-layer, resulting in predictions (i.e., RAP) at every spatial location for every individual. By computing the performance of RAP across all the spatial locations, one can determine which regions are informative for sex prediction. We applied the 100 models in the LEAP dataset, and then generated 100 predictions for every individual, as well as the corresponding RAP maps (spatial prediction for maleness probability). For each voxel of an individual, we used the median prediction from the 100 RAP maps to generate the ensemble prediction. We then compared the group differences of the maleness probability RAP maps.

The RAP generation steps for LEAP dataset are summarized as follows:

1. Every spatial location of each individual gets 100 predictions for the maleness probability.
2. The 100 predictions are summarized with the median value.
3. The process is repeated for all 681 individuals and every individual receives a median-RAP map.

### Associations with autism-associated, clinical features

To assess the clinical relevance of our findings, we followed results up in autistic individuals (males and females separately) within the deeply phenotyped LEAP sample. We ran two general linear model (GLM) for autistic males and females separately, with core clinical and cognitive measures associated with autism as independent variables while accounting for age and site. These core clinical and cognitive measures included: ADOS (CSS, CSS-SA, CSS-RRB), ADI-R (reciprocal social interaction, communication, RRB), SRS-2, RBS-R, SSP, Vineland Adaptive Behaviour Composite (ABC) Score and three subscales (communication, daily living skills, socialization). We predicted more autistic symptoms and cognitive difficulties with higher male prediction probability and thus computed one-tailed p-values. All results were FDR-corrected. Results revealed no significant associations.

### Associations with sex-differential, cognitive measures

To assess the relationship with sex-differential cognitive features, we followed results up in autistic individuals (males and females separately) within the deeply phenotyped LEAP sample. We ran two general linear model (GLM) for autistic males and females separately, with sex-differential, cognitive features as independent variables while accounting for age and site. These sex-differential, cognitive features included: the Reading the Mind in the Eyes Test (RMET), Autism Quotient (AQ), Empathy Quotient (EQ), and Systemizing Quotient (SQ). We predicted more male-typical scores (i.e., higher scores on the AQ and SQ, while lower scores on the RMET and EQ) with higher male prediction probability and thus computed one-tailed p-values. All results were FDR-corrected. These analyses yielded a nominally significant relationships between higher male sex prediction values with higher scores on the AQ in ASC-M (*t*=2.05, *P*=0.02, *q*=0.08), while a significant relationship with lower performance on the RMET test in ASC-F (*t*=-2.36, *P*=0.01, *q*=0.04, SF3A).

We further tested whether a shift towards maleness was also associated with more sex-differential cognitive measures in neurotypical individuals. We ran two general linear models – one in NT males and one in NT females separately with sex prediction values as dependent variables and age and site as covariates. We predicted that higher sex prediction probability values towards maleness would be associated with higher AQ and SQ scores, and lower EQ scores or lower accuracy on the RMET. We found no significant associations between greater predictive male probability and any of the cognitive measures.

### Cognitive decoding

We further investigated two specific RAP-imaging-t-maps (ASC-F vs. NT-F and ASC-M vs. NT-M) focussing on regions with *positive* t-values (as these represented where ASC-M and ASC-F showed higher male prediction probabilities). To explore the cognitive domains implicated in these, we used the Neurosynth Image Decoder (<http://neurosynth.org/decode/>; accessed on January 23^rd^ 2021) to visualize the top 100 terms most strongly associated with the two RAP-imaging-t-maps.

After excluding anatomical (e.g., fusiform gyrus) and redundant terms (e.g., object and objects), we visualized the remaining terms showing correlations with the imaging maps between *r*=0.2–0.06.

Results showed that the most common cognitive terms associated with the female RAP-t-maps were primarily related to face perception, visual processing and speech, whereas in males to motor and reward processing (Figure 2D). Based on these results we tested the association between the RAP-based sex prediction values (extracted from clusters with highest male prediction probabilities) and *a)* cognitive measures associated with motor and reward processes in males (i.e., ADI-R RRB, ADOS-2 CSS-RRB, RBS-R, measures of accuracy on monetary and social reward tasks) and *b)* cognitive measures related to face and communication in females (i.e., emotional face processing tasks (KDEF & Hariri), ADI-R communication and communicative adaptive functioning).

Next, in order to validate the specificity of RAP images relative to behavioural data, we cluster-corrected these two RAP-imaging-maps (Gaussian random field theory, voxel-level *Z*=3.1, cluster-forming *P*=0.001) to identify ROIs with highest male prediction probabilities in ASC-M and ASC-F. These were then binarized and used as masks (one male-specific and one female-specific mask) to extract the average RAP-values for each male/female autistic individual. These ROI-specific values were then submitted to two GLMs in autistic males and females separately to test the association between the RAP-based sex prediction values within the clusters with the highest male prediction probabilities and with different cognitive measures related to the cognitive domains identified in the previously described cognitive decoding analyses. Results were FDR-corrected. While we found no relationships in ASC-M, in ASC-F there was a significant association between predicted maleness and lower accuracy on the Karolinska Directed Emotional Faces task (*t*=-2.6, *P*=0.01, *q*=0.02) (SF3B).

### ADHD sample

To assess specificity of results with regards to autism, we selected an independent sample of individuals with attention-deficit/hyper-activity disorder (ADHD) and NT individuals. For this we combined the publicly available ADHD200 (47) (excluding the NeuroImage site collection) and the local NeuroImage (48) sample from Nijmegen and Utrecht (as the local sample is larger in size than the Nijmegen-NeuroImage sample, which is part of ADHD200). Given that the NeuroImage study is a sibling designed cohort, there were pairs of siblings who were either both control individuals or both individuals with a diagnosis of ADHD. We thus created a custom-code that randomly excluded one of the two sibling pairs, retaining only one member of the family (in both the control and ADHD category). Acquisition sites that provided less than four individuals (referred to as an individual’s imaging data) per sex/diagnostic group were excluded (in this case two sites, namely, University of Pittsburgh and Washington University in St. Louis from ADHD200). We further excluded participants with (clinically non-significant) reported atypicalities in the MR data (N=7) and excessive head motion or corrupted image quality (N=54). This resulted in a sample of 370 males with ADHD, 134 females with ADHD, 246 NT males and 220 NT females across six sites in total.

To account for a significant difference in age *(F*=1.7, *P*<0.001) between males and females with ADHD (mean age females=13.3, mean age males=14.2) and NT males and females (mean age females=12.5, mean age males=12.2), we used both auto-age matching implemented in our code (as described below) and we also manually matched individuals on age (mean age females with ADHD=13, mean age males with ADHD=13.3, mean age NT females=12.8, mean age NT males=12.6; *F*=1.97, *P*=0.12; after excluding 4 females with ADHD, 46 males with ADHD, 13 NT females, 21 NT males). This resulted in a final sample of 324 males with ADHD, 130 females with ADHD, 225 NT males and 208 NT females across six sites in total. Individuals were matched for age across sex/diagnostic groups and for FIQ within diagnostic groups (i.e., ADHD-M matched with ADHD-F; and NT-M matched with NT-F). For details, see ST2.

#### Sex prediction analyses in the ADHD sample

We applied the 100 fine-tuned models from ABIDE to the combined ADHD200 - Neuroimage sample the same way as described in the method sections in relation to the LEAP sample. More specifically, the models were applied to every individual in the ADHD testing dataset. Given the age distribution of the sample, we did not want results to be biased by older males and females with ADHD. Thus, we ran analyses in two samples: a) in the manually age-matched sample (as described above) and b) in a sample that was auto age-matched within the code when creating the sex- and clinically-balanced samples for testing. SF1D shows the resulting age distributions for the sex- and clinically-balanced sample when doing manual age-matching and SF1F shows the resulting age distributions for the sex- and clinically-balanced sample when doing auto age-matching within the code.

##### Manual age-matching (before the model application)

Results revealed that females with ADHD (72%) had similar sex prediction accuracies as males with ADHD (73%) (Cohen’s *d*=0.1, *P*=0.3), and slightly reduced compared to NT individuals (73%) (*d*=0.24, *P*=0.02), however this did not survive Bonferroni correction (*P*=0.05/3=0.017). Males with ADHD showed no differences in sex prediction accuracy from NT (d=0.05, *P*=0.6) (SF4A and ST5A).

##### Auto age-matching (within the model application)

Results revealed that females with ADHD (72%) had similar sex prediction accuracies as males with ADHD (71%) (Cohen’s *d*=0.14, *P*=0.2) and as NT (73%) (*d*=0.19, *P*=0.05). Males with ADHD showed slightly reduced sex prediction accuracies compared to NT (*d*=0.4, *P*=1.0e-04), however again this did not survive Bonferroni correction (*P*=0.05/3=0.017) (SF4B and ST5B).

### Comparison of autistic individuals with and without ADHD

In order to compare results in the LEAP sample with the ADHD sample results, we also computed classification accuracies across individuals with autism with and without a co-occurring diagnosis of ADHD (ADHD+ vs. ADHD-). Autistic individuals were sub-grouped based on DSM-5 criteria as specified above. This resulted in a sample of 120 autistic males with ADHD, 119 autistic males without ADHD (47 autistic males had missing values on the ADHD measure), 39 autistic females with ADHD and 61 autistic females without ADHD (9 autistic females had missing values on the ADHD measure). Note that 13 NT males and 11 NT females also scored above threshold on the ADHD rating scale, were however not excluded from analyses for consistency reasons. For accuracy comparisons between each diagnostic and sex group, the same number of individuals was sampled for each model, i.e. the smallest available number in a certain group, which was 39 (ASC-F) in this case.

Results revealed that the previously reported pattern was present across both autistic males and females with and without a co-occurring diagnosis of ADHD (ST6). We found that the sex prediction accuracy in autistic females with ADHD (71%) was on average lower than that in autistic males with ADHD (87%) (Cohen’s *d*=1, *P*=1.4-e16) and less than that in NT (80%) (*d*=-0.7, *P*=1.5e-10). For the male individuals, sex prediction accuracy in autistic males with ADHD was on average better than in NT (*d*=0.7, *P*=2.1e-10) (SF5A). Sex prediction accuracy in autistic females without ADHD (75%) was on average less than that in autistic males without ADHD (86%) (*d*=-0.7, *P*=9.8-e11) and that in NT individuals (*d*=-0.5, *P*=1.9-e05). For the male individuals, performance in autistic males without ADHD was on average better than that in NT individuals (*d*=-0.6, *P*=6.9-e09) (SF5B). Autistic males with ADHD did not differ from autistic males without ADHD (*d*=-0.03, *P*=0.8), whereas autistic females with ADHD had slightly lower sex prediction accuracies than autistic females without ADHD (*d*=-0.02, *P*=0.03) (however not surviving Bonferroni correction *P*=0.05/2=0.025) (SF5C).

### Sensitivity / Control analyses

All method-based control analyses were done in the ABIDE validation sample, while sample-specific control analyses were carried out in the LEAP sample.

#### Age

Despite the absence of significant age differences between the sex and diagnostic groups, we still also included an auto age-matching algorithm into our model code to confirm results when the four samples were perfectly matched within the prediction models. The resulting age distribution of the sex- and clinically-balanced LEAP sample (N=340) can be seen in SF1E**.** Results remained unchanged to those from the original pipeline (see SF6A and ST7A).

Further, our finding of reduced and superior prediction performance in autistic females and males, respectively, is most pronounced in childhood and decreases throughout development to young adulthood. Only in the last age bin of around 20-30, we do not observe classification differences across groups. We carefully reviewed the possibility of a trivial technical explanation: although we have more adult individuals in the pre-training step in UKB, the classifier depends mostly on the fine-tuning (training) stage where we retrain all learnable parameters (44). Since the individual age distribution in this final fine-tuned dataset (ABIDE) peaks at around 12-years, any accuracy bias originating from the sample distribution should be around the younger rather than older individuals in our case.

#### Total intracranial volume

All the imaging inputs were controlled for brain volume through the 12 degree-of-freedom linear registration step in the preprocessing pipeline. Still, we double-checked whether there were any significant group differences between NT-F and ASC-F and between NT-M and ASC-M in total intracranial volume that might have influenced observed sex prediction patterns. First, we compared Freesurfer derived total intracranial volume estimates (before linear registration) across the groups. Two t-tests between NT-F and ASC-F (*t*=-0.74, *P*=0.46) and between NT-M and ASC-M (*t*=-1.5, *P*=0.12) revealed no significant group differences. Next, we included an automatic brain-volume-matching algorithm on top of the age-matching algorithm to ensure balanced age and brain volume across sex and diagnostic groups. More specifically, individuals were divided into age bins spanning five years each. Within each age bin, individuals were ranked by their total intracranial volume, and divided into ten sub-groups. Hence, individuals in each sub-group were within the 10% brain volume rankings across the population within each age bin. We then sampled the same number of individuals from each sex-clinical label for each subgroup, which ensured that the final population was balanced across age and volume. Based on this, we compared the prediction differences between each sex and diagnostic groups again. Results remained unchanged (SF6B and ST7B).

#### Model choice

##### Autism probability as a function of sex prediction probability

Previously, we established that different diagnostic groups give rise to differential sex prediction accuracies. Here, by reversing the model, we confirm that conversely the different accuracies of sex prediction give rise to a stratification along the diagnostic groupings – even though using non-linear models (where compared to linear models x [dependent variable] and y [independent variable] cannot simply be interchanged). To do this, we investigated whether the probability of having autism was related to the sex predictions (49). We created eight probability bins in steps of 0.125 and determined the sample probability in each bin by dividing the total number of ASC-F (ASC-M) by the total number of females (males) in each bin and ran Spearman’s correlations between the sample probability and the predictive sex probability. Next, we subdivided autistic individuals into a) correctly classified females (*P*<0.5) / males (*P*>0.5) and b) misclassified females (*P*>0.5) / males (*P*<0.5). Within each sex, we compared the proportions of having autism as a function of being correctly or incorrectly classified.

In ASC-F the sample probability of autism increased with increasing predictive probabilities for being classified as male (*rho*=-0.89, *P*<0.01), while there was no such relationship in ASC-M (*rho*=0.6, *P*=0.1; SF6C, left). ASC-F misclassified as male were significantly more likely to have an autism diagnosis than autistic females correctly classified as female (*P*=.82 vs. *P*=.52; χ^2^=3.96, *P*=0.02). There was no such relationship in ASC-M (*P*=.62 vs. *P*=0.56; *χ^2^*=1.59, *P*=0.1).

##### Principal component analysis with logistic regression

To check model-dependence of our results in ABIDE, we applied an alternative, but simpler method. Following the same train/validation pattern as we applied within the CNN method, we ran the following principal component analysis (PCA) based experiments 100 times: we used 75 principal components explaining most of the variance of the imaging data, and subsequently trained a logistic regression classifier for sex classification. Results showed the same patterns as we observed before: reduced sex prediction accuracy in autistic females compared to autistic males (Cohen’s *d*=1.4, *P*=2.8e-25) and compared to NT (*d*=1.4, *P*=8.4e-25), while increased sex prediction accuracy in autistic males compared to NT (*d*=0.7, *P*=3.6e-10), however with significantly reduced prediction performance (SF6C, right and ST7C). In this regard, we want to point out that PCA combined with logistic regression only takes into account features that contain very limited information (via PCA decomposition), which often reflects the global features in brains. Thus, it is not surprising that the sex prediction accuracy drops considerably compared to the classification results based on our CNN, model which learned non-linear features and classifiers.

#### RAP validation (mask out salient regions)

To verify the RAP method in ABIDE, we decided to compare sex prediction results when only using the most salient brain regions versus excluding the most salient brain regions in the RAP maps. For this, we generated two masks: one ASC-sensitive and the other ASC-insensitive mask. To obtain the ASC-sensitive mask, we thresholded the two RAP t-maps (map-A: ASC-F vs. NT-F; map-B: ASC-M vs. NT-M) to retain only the 25% highest values within each. Next, we combined the two thresholded maps using logical union (map-C = map-A ∪ map-B), and smoothed and dilated (50) the resulting map-C. The ASC-insensitive mask was the difference between the MNI mask and the ASC-sensitive mask. Both resulting masks contained about 50% of voxels within the MNI brain mask and were thus comparable in size. Next, the RAP method was re-run twice, once within the ASC-sensitive mask and once within the ASC-insensitive mask and prediction accuracies were computed as specified earlier.

Results revealed that the previously observed sex prediction accuracy differences were more pronounced when applying the ASC-sensitive than the ASC-insensitive mask (ST7D). Specifically, using the ASC-sensitive RAP mask, sex prediction accuracies were significantly lower in autistic females compared to autistic males (Cohen’s *d*=1.7, *P*=3.8-e16) and to NT (*d*=1.4, *P*=1.6e-13). Autistic males also showed reduced sex prediction performance compared to NT individuals (*d*=1.1, *P*=6.0e-10) (SF6D, left). Using the ASC-insensitive mask, similar patterns albeit with smaller effect sizes emerged (ASC-F vs. ASC-M: *d*=0.6, *P*=5.6e-05; ASC-F vs. NT: *d*=0.3, *P*=2.6e-02; ASC-M vs. NT: *d*=0.5, *P*=3.9e-04; SF6D, right).

#### Comparison of variance in sex prediction accuracies across groups

We compared the ratios of variances of sex prediction accuracies across the groups using the var.test function in R. NT-M had significantly higher variance across prediction accuracies compared to NT-F (*F*=0.15, *P*<0.001). There was also a sex differences in variance in autistic individuals, however with ASC-M having lower variance compared to ASC-F (*F*=0.4, *P*<0.001). ASC-F had significantly higher prediction variance compared to NT-F (*F*=2.22, *P*<0.001), while there was no significant difference between ASC-M and NT-M (*F*=0.82, *P*=0.12).

Previous research shows that the male population displays higher intrinsic variability than females (51). Here we also see higher prediction variability across males than across females. We exclude however the possibility that sex prediction accuracy differences are driven by variance differences: if sample variance was the same across NT-M and NT-F, we would expect a) improved classification accuracies for both NT-M and NT-F (as our CNN was trained to predict maleness), b) even worse classification accuracies for autistic females. Thus, homogeneity of NT-sample variance would even further accentuate our results in autistic females, however, strikingly, our findings of reduced classification accuracies in autistic females hold even at the background of differences in NT-variance.

### Comparison of autistic individuals with high and low prediction accuracies

We applied a quartile split on the prediction probability values in autistic males and females to subdivide them into highly misclassified (lower 25% - ‘ASC-M/F low’) and highly correctly classified (upper 25% - ‘ASC-M/F high’) individuals. These two classes of autistic males/females were compared by computing t-maps (ASC-M/F low vs. ASC-M/F high) based on their RAP maps which were subsequently cluster-corrected (using Gaussian random field [GRF] theory, voxel-level Z=3.1, cluster-forming *P*=0.001) to identify the most differentiating regions of interest (ROIs). When comparing the top 25% with the bottom 25% of autistic males and females on their RAP contrast maps, the most differentiating regions between highly misclassified and highly correctly classified autistic females were in cerebellum, left thalamus, superior frontal gyrus and frontal medial cortex in grey matter, and left retrolenticular part of internal capsule, posterior thalamic radiation, right anterior corona radiata and the splenium of corpus callosum in white matter (Figure 7A). The most differentiating regions between highly misclassified and highly correctly classified autistic males were in bilateral superior parietal lobule, bilateral supplementary motor cortex, precuneus, superior lateral occipital cortex, paracingulate gyrus, frontal medial cortex and frontal pole in grey matter, and bilateral superior cerebellar peduncle and right superior longitudinal fasciculus in white matter (SF7B).

### Gene expression decoding

We next wanted to address the question of what the potential sources of masculinization were by tapping into the underlying genomic mechanisms.

First, RAP-imaging-t-maps were uploaded on neurovault (ASC-F vs. NT-F and ASC-M vs. NT-M and NT-F vs. NT-M as reference; see

(<https://identifiers.org/neurovault.collection:9354>). We then used the gene expression dataset from the Allen Human Brain Gene Expression atlas (AHBA)^25,26^ which includes samples from six post-mortem brains (3 Caucasian, 2 African, 1 Hispanic; 1 female) ages 24-57 years. This limited sample size and large variability across age, sex and ethnicity can impact the transcriptional patterns. Previous studies addressing this have shown that results were not driven by one single donor and generalizable beyond the donor brains in the AHBA after rigorously testing the effect of donor selection by running leave-one-donor-out analyses (52) and using Bayesian random effects analysis which fitted a hierarchical Bayesian regression model using Markov Chain Monte Carlo sampling (53). We employed the gene expression decoding functionality (53) integrated in Neurosynth(54) ([https://neurosynth.org](https://neurosynth.org/)) and NeuroVault (55) ([https://neurovault.org](https://neurovault.org/)). This performs in detail the following steps: it first uses the Allen Brain Atlas REST API to download the gene expression data, extracts the MNI coordinates for each sampling site, draws a spherical ROI (4mm) and extracts the average values of the statistical maps within each ROI. In a fixed effects analysis, the resulting vector of values can next be correlated with the normalized gene expression values (20,787 protein decoding genes from all donors pooled together) to see how similar they are. Next, an approximate random effects analysis calculates the slope of best linear fit for each donor (individually fitted regression lines) and performs a one-sample t-test on those estimates to test how consistent the relations between the gene expression and evaluated map values are and identifies genes whose spatial expression patterns are consistently across the six donor brains highly similar to the evaluated maps. The resulting list of genes was next corrected for multiple comparisons and only genes surviving FDR *P*<0.05 were retained.

### Gene classes in enrichment analyses

After obtaining this list of genes that were highly expressed in spatial patterns throughout the brain and similar to the two sex-differentiation RAP maps, we went on to test the overlap (i.e. enrichment) of our gene lists with a set of relevant classes of genes. We opted for three different gene classes: **1)** **autism-associated genes** to investigate whether the genetic likelihood for autism overlaps with the male neurophenotype; **2)** genes acting in **prenatal** development as events during the embryonic period have long-lasting effects on both sexual differentiation and susceptibility for neurodevelopmental conditions (prenatal cell types (56)); and **3)** **sex-differentially expressed** **genes** in prenatal development and genes **differentially regulated by prenatal sex steroids**.

#### Autism associated genes

##### Structural genetic variants

Structural genetic variants included common genetic variants and de novo mutations. Common genetic risk variants were obtained from a large-scale genome-wide association study by Grove et al. (57) SNP based P-values were converted to gene-based P-values using a hg19 genome build using MAGMA (58) which accounts for linkage disequilibrium between SNPs when calculating gene-based P-values. The identified list of genes was subsequently Bonferroni corrected (ASC commonRV). Genes harbouring rare, de novo variants associated with autism were identified by Satterstrom et al. (59) doing whole-exome sequencing of 35,584 samples (11,986 with autism). A total of 102 rare, de novo protein truncating genes associated with autism (FDR adjusted *P*-value < 0.1) were identified (ASC dnPTV). Parikshak et al. (60) identified fetal gene co-expression modules associated with autism genes that have highly similar expression patterns during cortical development(60). Authors used RNA sequencing on gene expression data from BrainSpan whole-genome transcriptomic data and ran weighted gene co-expression network analysis. Among the five co-expression modules associated with different forms of autism risk, we selected modules 2 and 3 as transcriptional regulators were enriched for rare genetic variants in early fetal development (M2 and M3).

##### Transcriptionally dysregulated genes

To capture genetic downstream effects, we selected a range of transcriptionally dysregulated genes in autism. Gandal et al. (61) identified differentially expressed genes from an analysis of RNA sequencing data from the PsychENCODE Consortium, using autism post-mortem frontal and temporal cortex tissue (FDR adjusted *P*-value < 0.05) (865 downregulated: ASC DE Downreg, 746 upregulated: ASC DE Upreg)(61).

Parikshak et al.(62) performed rRNA-depleted RNA sequencing using post-mortem frontal and temporal cortical tissue samples from 48 autistic individuals and 49 control individuals. Doing weighted gene co-expression network analysis, they identified transcriptionally dysregulated co-expression modules in autism. Among these, the downregulated modules were enriched in synaptic function and neuronal genes, whereas upregulated modules were enriched in genes associated with inflammatory pathways and glial functions (764 downregulated: ASC CTX Downreg CoExpMods, 1111 upregulated: ASC CTX Upreg CoExpMods) (62).

Performing single-nucleus RNA sequencing analysis of cortical tissue of individuals with autism, Velmeshev et al. (63) analysed the transcriptomes of single brain cells. They generated 104,559 single-nuclei gene expression profiles (52,556 from control individuals and 52,003 from autistic individuals) and performed unbiased clustering of nuclear profiles and annotated clusters based on expression of known cell type markers. This way they identified genes differentially expressed in autism in specific cell types (ASC Excitatory, ASC Inhibitory, ASC Microglia, ASC Oligodendrocyte, ASC Astrocyte, ASC Endothelial).

#### Prenatal cell types

This gene list was based on the study by Polioudakis et al. (56) who performed RNA sequencing on 40,000 cells and created a single-cell gene expression atlas of the developing, mid-gestation (gestation weeks 17 to 18) human neocortex. This resulted in the identification of 16 transcriptionally distinct cell groups which clustered by known major biological cell types at this stage of development, including: endothelia (E), excitatory neurons (EN; migrating excitatory, maturing excitatory, maturing excitatory upper enriched, excitatory deep layer 1, excitatory deep layer 2), interneurons (IN; interneuron MGE and interneuron CGE), intermediate progenitors (IM), microglia (M), cycling progenitors (MN; cycling progenitor S-phase and cycling progenitors G2M phase), oligodendrocyte precursors (OPC), pericyte (P), and radial glia (RG; ventricular radial glia and outer radial glia).

#### Sex-differentially expressed genes

We further surveyed sex-differentially, prenatally expressed genes. For this, we used the same sex-differential gene array expression data from prenatal samples as Werling et al. (64). This was based on a study by Kang et al. (65) who analyzed exon array data from individuals between 16 and 22 post-conception weeks from frontal, temporal and parietal cortex. For genes differentially regulated by sex hormones, we used gene lists from two studies: 1) Quartier et al. (66) conducted an RNA sequencing study in which embryonic neural stem cells were treated with 100nM of dihydrotestosterone (DHT), a potent, non-aromatizable androgen. We used the same list of genes differentially expressed by DHT as Lombardo et al. (67, 68). 2) Willsey et al. (69) conducted RNA sequencing and identified differentially expressed genes in Xenopus whole brain following 17-β-estradiol treatment. We used the list of genes differentially expressed by estrogen.

### Enrichment analyses

Analyses examining enrichment were done with a custom code written by MVL (<https://github.com/mvlombardo/utils/blob/master/genelistOverlap.R>) computing the enrichment odds ratios and hypergeometric p-values for all enrichment hypergeometric tests based on the sum(dhyper) function in R. The background set size for enrichment analyses was set to the number protein decoding genes considered in the Neurosynth Gene Expression Decoding analyses (i.e., 20,787). To avoid biasing our findings towards genes expressed in brain, we conducted another enrichment analysis using a more conservative list of 16,906 background genes based on real estimates of genes expressed in cortical tissue (70) (see SF8). Only comparison with FDR *P*<0.05 were interpreted further.

8.	Del Barrio V: Diagnostic and statistical manual of mental disorders, inThe Curated Reference Collection in Neuroscience and Biobehavioral Psychology. 2016

9.	American Psychiatric Association: Diagnostic and statistical manual of mental disorders: DSM-IV-TR (text revision). 2000

10.	American Psychiatric Association: Diagnostic and Statistical Manual of Mental Disorders 1. American Psychiatric Association. 2013

11.	World Health Organization: The ICD-10 classification of mental and behavioural disorders: Diagnostic criteria for research. 1993

12.	Wechsler D: Wechsler Abbreviated Scale of Intelligence–Second Edition (WASI‐II). San Antonio, TX, NCS Pearson., 2011

13.	Wechsler D: Wechsler Intelligence Scale for Children (3rd ed.). San Antonio, TX, Psychological Corporation, 1991

14.	Wechsler D: Wechsler Intelligence Scale for Children (4th ed.). San Antonio, TX, Psychological Corporation, 2003

15.	Wechsler D: Wechsler Adult Intelligence Scale (3rd ed.). San Antonio, TX, The Psychological Corporation, 1997

16.	Wechsler D: Wechsler Adult Intelligence Scale–Fourth Edition. San Antonio, TX, Pearson, 2008

17.	Hus V, Gotham K, Lord C: Standardizing ADOS domain scores: Separating severity of social affect and restricted and repetitive behaviors. Journal of Autism and Developmental Disorders 2014; 44:2400–2412

18.	Gotham K, Risi S, Pickles A, et al.: The autism diagnostic observation schedule: Revised algorithms for improved diagnostic validity. Journal of Autism and Developmental Disorders 2007; 37

19.	Constantino JN, Gruber &: Social Responsiveness Scale-Second Edition (SRS-2). Journal of Psychoeducational Assessment 2014;

20.	Bodfish JW, Symons FJ, Parker DE, et al.: Repetitive Behavior Scale–Revised. PsycTESTS® 2000;

21.	Tomchek SD, Dunn W: Sensory processing in children with and without autism: a comparative study using the short sensory profile. American Journal of Occupational Therapy 2007; 61

22.	Sparrow SS, Cicchetti D V., Balla DA: The Vineland Adaptive Behavior Scales (2nd ed), inMajor psychological assessment instruments. 2005

28.	Baron-Cohen S, Wheelwright S: The empathy quotient: An investigation of adults with asperger syndrome or high functioning autism, and normal sex differences. Journal of Autism and Developmental Disorders 2004; 34

29.	Baron-Cohen S, Wheelwright S, Hill J, et al.: The “Reading the Mind in the Eyes” Test revised version: A study with normal adults, and adults with Asperger syndrome or high-functioning autism. Journal of Child Psychology and Psychiatry and Allied Disciplines 2001; 42

30.	Baumeister S, Moessnang C, Bast N, et al.: Attenuated Anticipation of Social and Monetary Rewards in Autism Spectrum Disorders. bioRxiv 2020;

31.	Hariri AR, Tessitore A, Mattay VS, et al.: The amygdala response to emotional stimuli: A comparison of faces and scenes. NeuroImage 2002; 17

32.	Goeleven E, De Raedt R, Leyman L, et al.: The Karolinska directed emotional faces: A validation study. Cognition and Emotion 2008; 22

33.	Lundqvist D, Flykt A, Ohman A: The Karolinska directed emotional faces (KDEF). CD ROM from Department of Clinical Neuroscience, Psychology section, Karolinska Institutet 1998;

34.	Sucksmith E, Allison C, Baron-Cohen S, et al.: Empathy and emotion recognition in people with autism, first-degree relatives, and controls. Neuropsychologia 2013; 51

35.	Smith SM: Fast robust automated brain extraction. Human Brain Mapping 2002; 17:143–155

36.	Peng H, Gong W, Beckmann CF, et al.: Accurate brain age prediction with lightweight deep neural networks. Medical Image Analysis 2021; 68

37.	Simonyan K, Zisserman A: Very deep convolutional networks for large-scale image recognition, in3rd International Conference on Learning Representations, ICLR 2015 - Conference Track Proceedings. 2015

38.	Shelhamer E, Long J, Darrell T: Fully Convolutional Networks for Semantic Segmentation. IEEE Transactions on Pattern Analysis and Machine Intelligence 2017; 39

39.	Ioffe S, Szegedy C: Batch normalization: Accelerating deep network training by reducing internal covariate shift, in32nd International Conference on Machine Learning, ICML 2015. 2015

40.	Lecun Y, Bengio Y, Hinton G: Deep learning. Nature 2015; 521:436–444

41.	Srivastava N, Hinton G, Krizhevsky A, et al.: Dropout: A simple way to prevent neural networks from overfitting. Journal of Machine Learning Research 2014; 15

42.	Paszke A, Gross S, Massa F, et al.: PyTorch: An imperative style, high-performance deep learning library, inAdvances in Neural Information Processing Systems. 2019

43.	Weiss K, Khoshgoftaar TM, Wang DD: A survey of transfer learning. Journal of Big Data 2016; 3

44.	Holderrieth P, Weikang G, Smith SM, et al.: Transfer learning works in neuroimaging via feature re-use, inMIDL 2021 Conference. 2021

45.	Pan SJ, Yang Q: A survey on transfer learning. IEEE Transactions on Knowledge and Data Engineering 2010; 22

46.	Raghu M, Zhang C, Kleinberg J, et al.: Transfusion: Understanding transfer learning for medical imaging. arXiv 2019;

47.	Milham PM, Damien F, Maarten M, et al.: The ADHD-200 Consortium: A model to advance the translational potential of neuroimaging in clinical neuroscience. Frontiers in Systems Neuroscience 2012;

48.	von Rhein D, Mennes M, van Ewijk H, et al.: The NeuroIMAGE study: a prospective phenotypic, cognitive, genetic and MRI study in children with attention-deficit/hyperactivity disorder. Design and descriptives. European Child and Adolescent Psychiatry 2015; 24:265–281

49.	Ecker C: Notice of Retraction and Replacement: Ecker et al. Association between the probability of autism spectrum disorder and normative sex-related phenotypic diversity in brain structure. JAMA Psychiatry. 2017;74(4):329-338. JAMA Psychiatry 2019; 76:549–550

50.	Gonzalez R, Woods R: Digital image processing (answers). 2002

64.	Werling DM, Parikshak NN, Geschwind DH: Gene expression in human brain implicates sexually dimorphic pathways in autism spectrum disorders. Nature Communications 2016; 7

### Supplemental Figures

#### **SF1 - Age distributions per sample and model step**

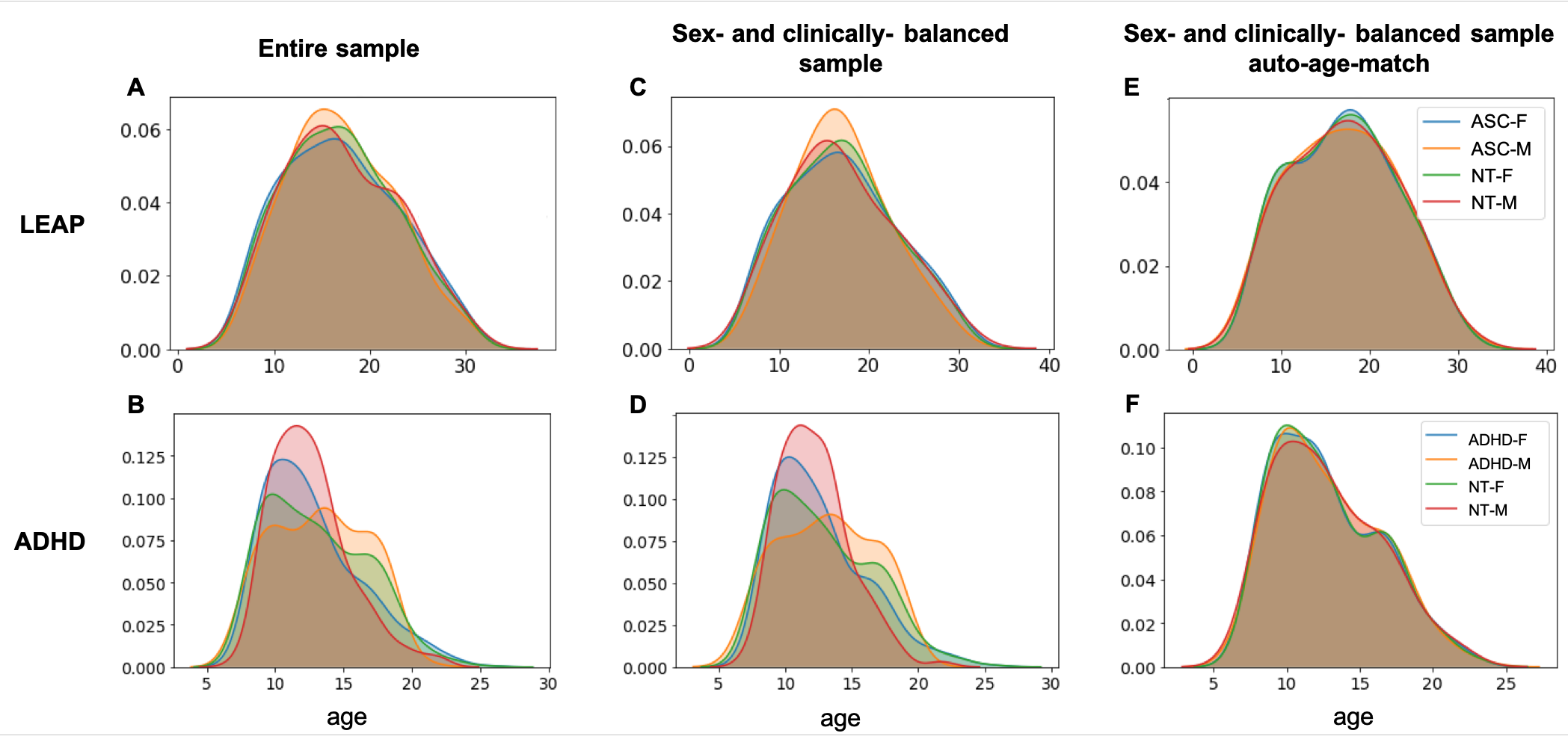

**SF1. A**) The age range in the entire LEAP sample comprised 6 to 31 years of age. For details see Table 1**. B**) The age range in the entire ADHD sample comprised 7 to 29 years of age. For details see Table S2. **C**) Age distribution in the sex- and clinically-balanced LEAP sample. **D**) Age distribution in the sex- and clinically-balanced ADHD sample when applying age-matching *before* the model application. **E**) Age distribution in the sex- and clinically-balanced LEAP sample when applying age-matching *within* the model application. **F**) Age distribution in the sex- and clinically-balanced ADHD sample when applying age-matching *within* the model application. As described in the main manuscript, ‘sex- and clinically-balanced’ refers to randomly sampling the cohort 100 times for training and validation to get balanced, un-biased numbers of individuals in each diagnostic (ASC/ADHD/NT) and sex group (M/F) across all sites. For further details, see Table S4. **Abbreviations**: ASC-F=autistic females, ASC-M=autistic males, NT-F= neurotypical females, NT-M= neurotypical males, ADHD= attention-deficit/hyperactivity disorder, ADHD-F= females with ADHD, ADHD-M=males with ADHD.

#### **SF2 - Simple Fully Convolutional Network**

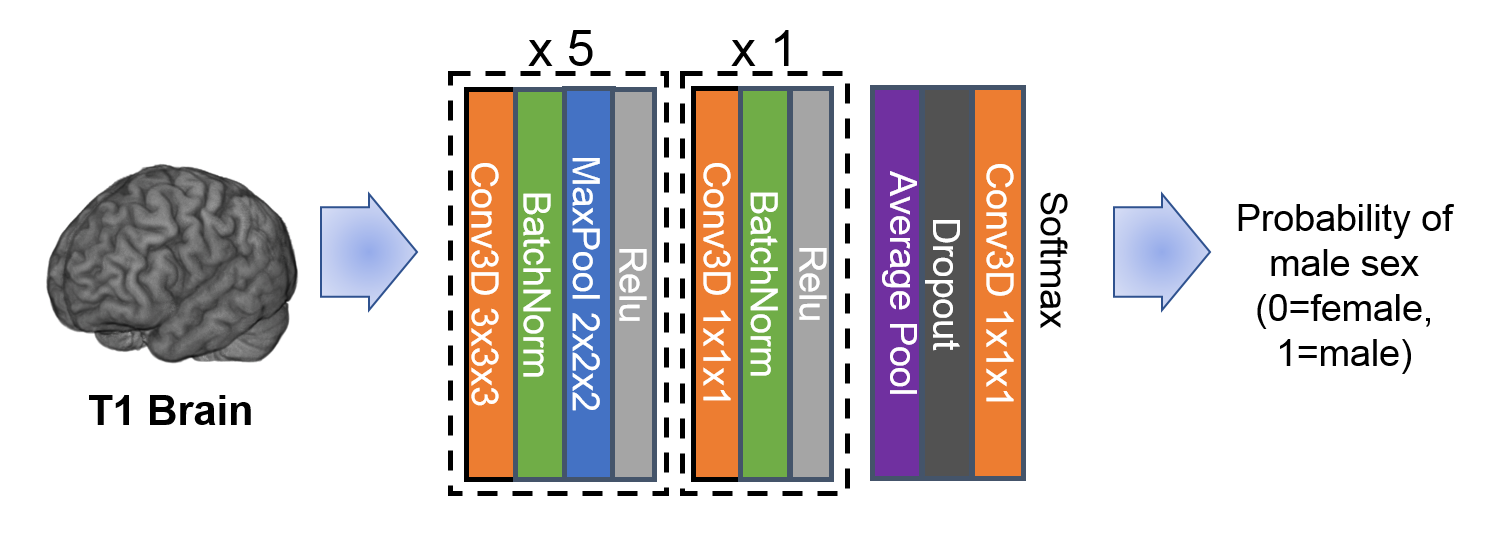

**SF2.** The deep neural network architecture used in this work: simple fully convolutional neural network (SFCN). The network takes linearly registered MRI brain 3D images, and consists of five Conv3D[3x3x3]-BatchNorm-MaxPool[2x2x2]-ReLU building blocks, and one Conv3D[1x1x1]-BatchNorm-ReLU building block, and then the average pooling and the final fully connected (FC) layer (effectively implemented as Cov3D[1x1x1]). The FC-layer outputs are two values, converted through a softmax layer to create the final outputs representing the ‘probability’ of sex prediction (i.e., predictive confidence).

#### **SF3 – Cognitive associations**

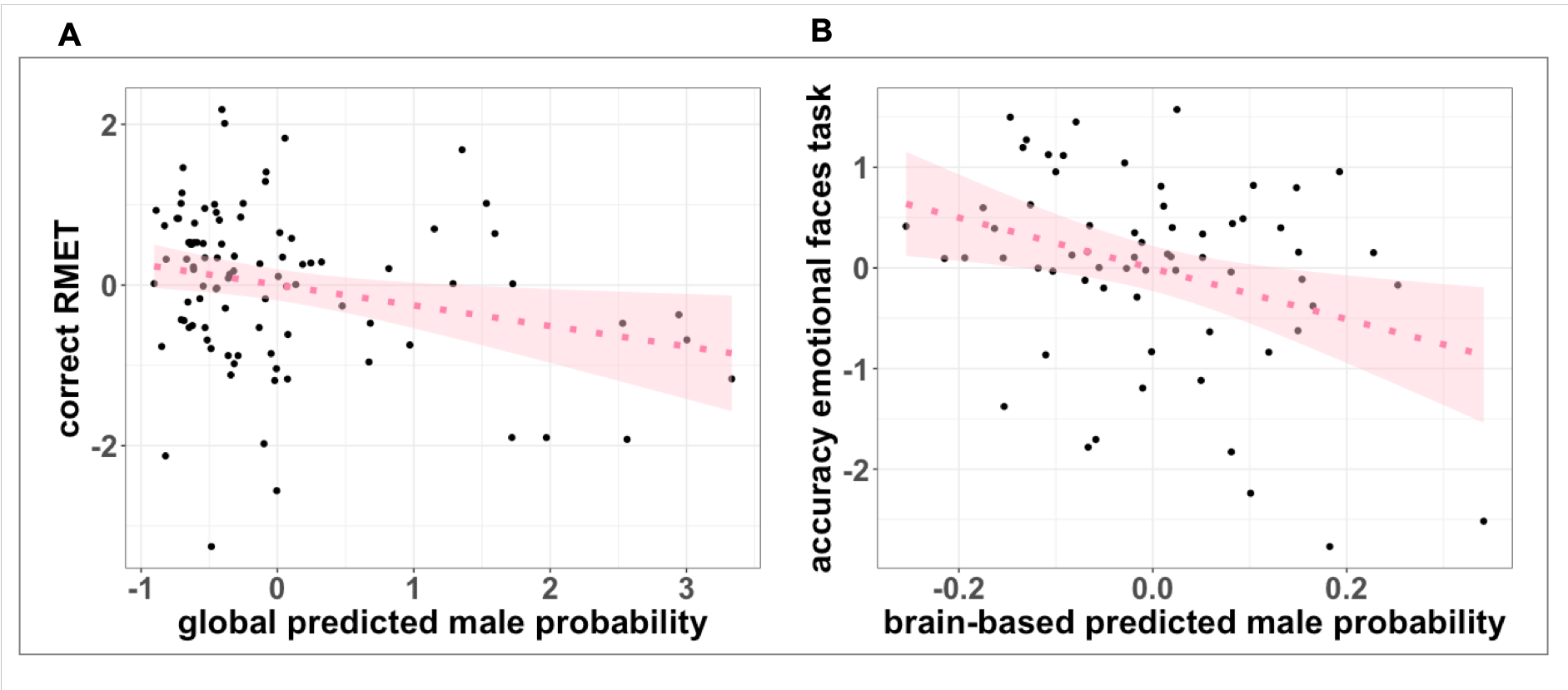

**SF3.** Association between predicted male probability (i.e., predictive confidence) and sex-differential cognitive features. **A**) Association between the global sex prediction probabilities and correct performance on the Reading the Mind in the Eyes task (RMET) in autistic females. **B**) Association between values extracted from the female RAP-imaging-t-maps (brain-based sex prediction values) and emotional face recognition difficulties (KDEF) in autistic values. Plotted values are Z-standardized.

#### **SF4 – ADHD sample prediction accuracies**

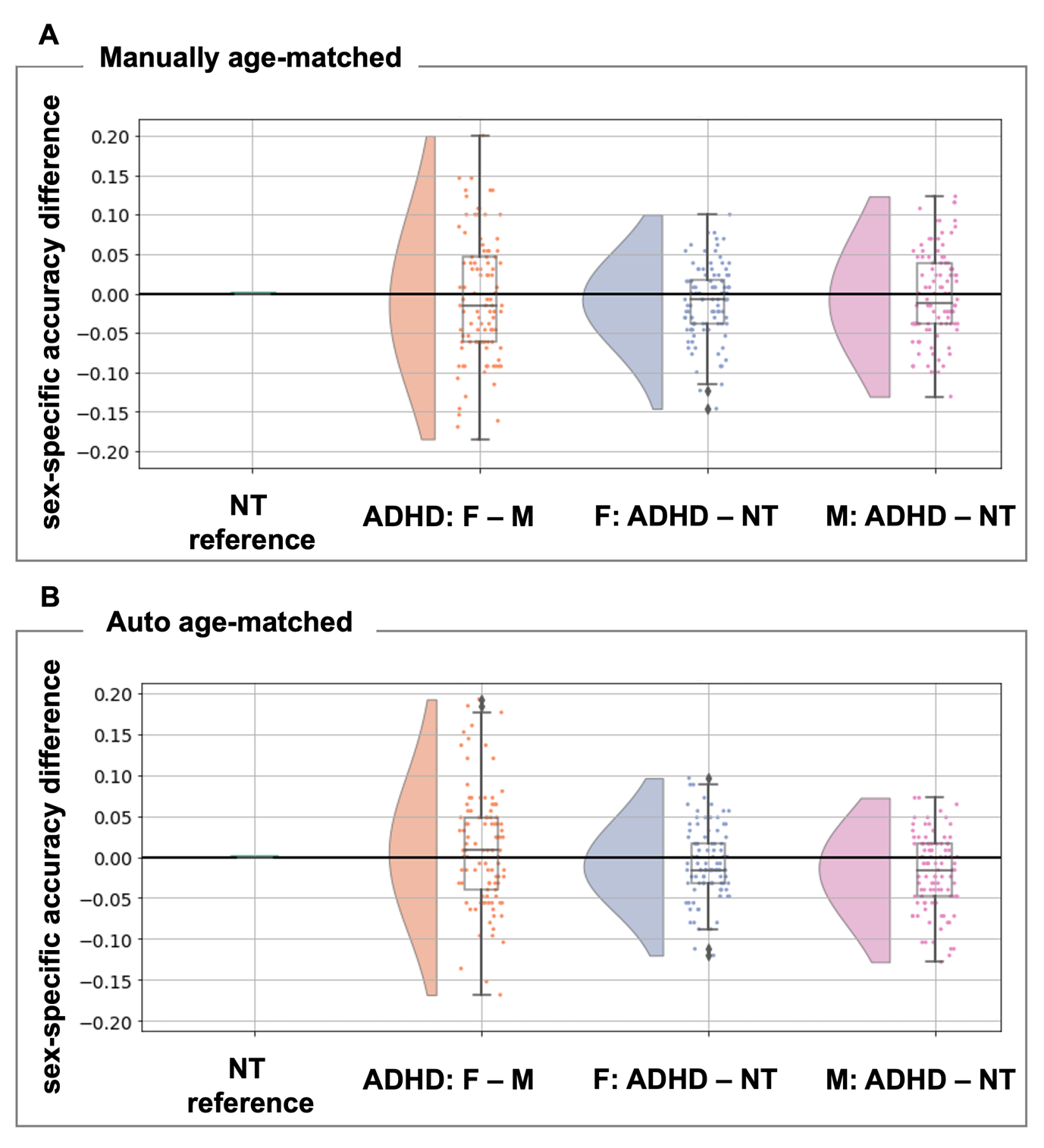

**SF4.** Comparison of sex prediction model performance across diagnostic and sex groups. We compare sex prediction accuracy differences between males and females with ADHD (ADHD: F – M), females with ADHD and neurotypical females (F: ADHD – NT) and males with ADHD and neurotypical males (M: ADHD – NT) in both the clinically- and sex-balanced sample matched for age before model application (**A**) and in the clinically- and sex-balanced sample matched for age within the model application (**B)**. Each dot represents one model out of 100 models in total. Negative values mean the model performs worse in the first sex/diagnostic group. Abbreviations: NT=neurotypical, ADHD=attention-deficit/hyperactivity disorder, F=females, M=males.

#### **SF5 – Comparison of autistic individuals with and without ADHD**

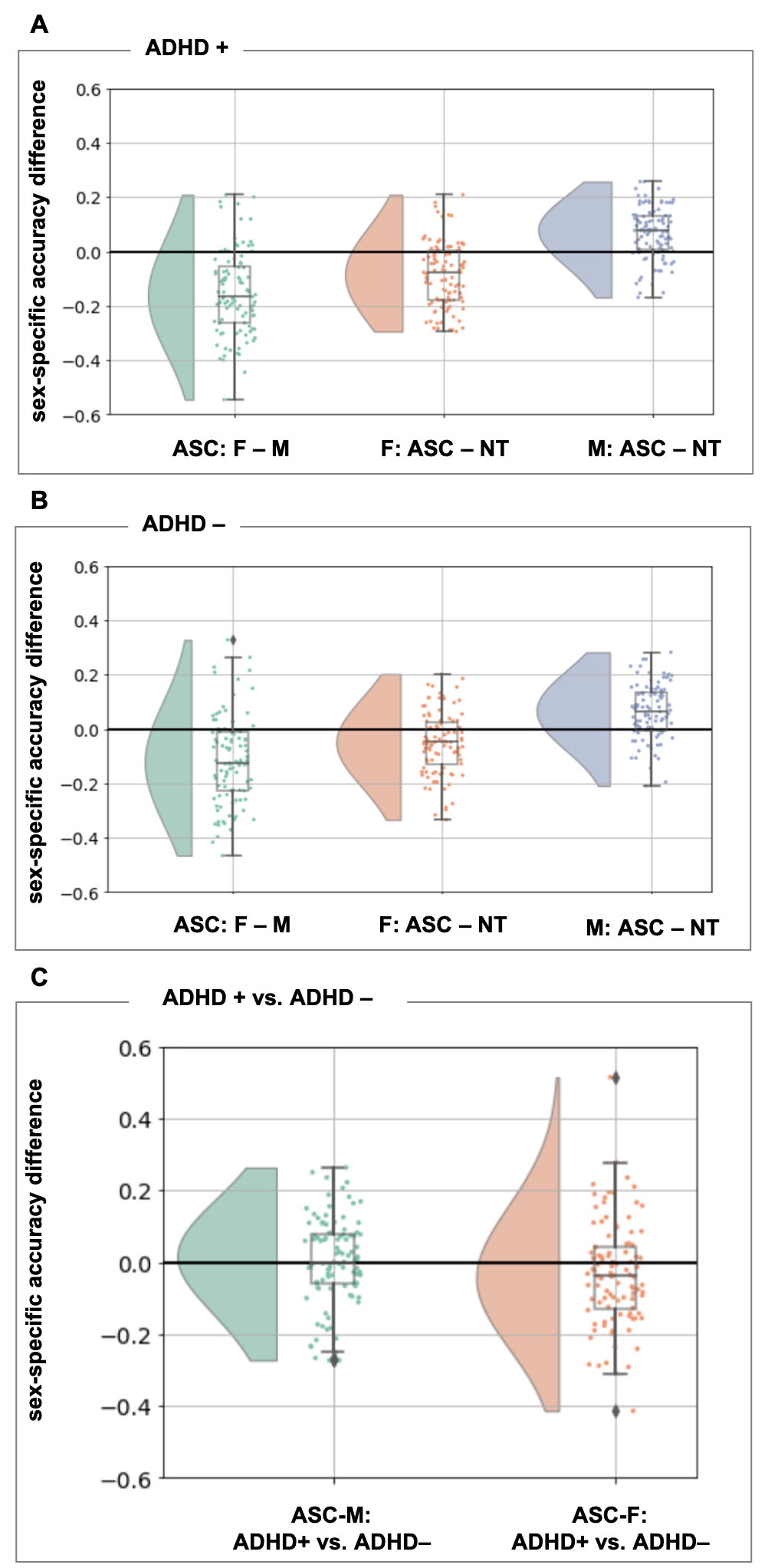

**SF5.** Comparison of sex prediction model performance across diagnostic and sex groups. We compare sex prediction accuracy differences between autistic males and autistic females (ASC: F – M), autistic females and neurotypical females (F: ASC – NT) and autistic males and neurotypical males (M: ASC – NT) in LEAP including either autistic individuals with co-occurring ADHD (**A**) or autistic individuals without co-occurring ADHD (**B**). In **C**) we compare sex prediction model performance between autistic males with and without ADHD and autistic females with and without ADHD. Each dot represents one model out of 100 models in total. Negative values mean the model performs worse in the first sex/diagnostic group. Abbreviations: ADHD+=autistic individuals with co-occurring ADHD, ADHD– = autistic individuals without co-occurring ADHD, NT=neurotypical, ASC=autism, ADHD=attention-deficit/hyperactivity disorder, F=females, M=males.

#### **SF6 – Sensitivity and control analyses**

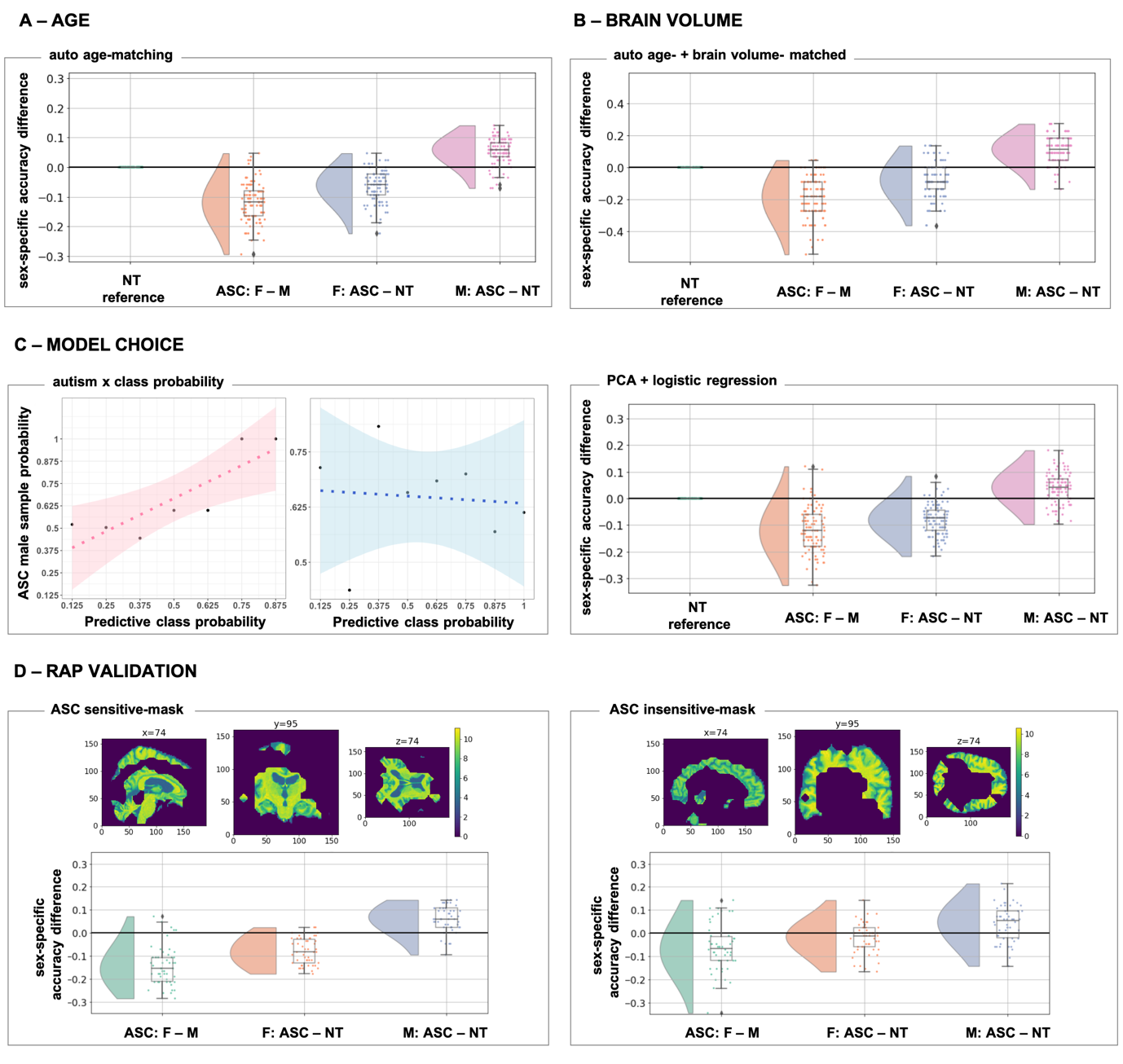

**SF6.** Overview of all sensitivity and control analyses. **A**) AGE. We included an auto age-matching algorithm into our model code to confirm results when the four samples were perfectly matched within the sex prediction models for age. We plot the sex prediction model performance across diagnostic and sex groups comparing sex prediction accuracy differences between autistic males and autistic females (ASC: F – M), autistic females and neurotypical females (F: ASC – NT) and autistic males and neurotypical males (M: ASC – NT) in LEAP. Each dot represents one model out of 100 models in total. Negative values mean the model performs worse in the first sex/diagnostic group. **B**) TOTAL INTRACRANIAL VOLUME. We extended the auto-age-matching algorithm to also match individuals on total intracranial volume. **C**) MODEL CHOICE. Left: Association between autism likelihood (sample probability) and predictive sex class probabilities. There is an association in autistic females (pink), however not in males (blue). Right: Principal component analysis combined with logistic regression: we used 75 principal components explaining most of the variance of the imaging data, and subsequently trained a logistic regression classifier for sex classification. **D**) RAP VALIDATION. We validated the RAP method in ABIDE by comparing sex prediction accuracy results when only using the most salient brain regions (left panel: ‘ASC-sensitive mask’) versus excluding the most salient brain regions in the RAP maps (right panel: ‘ASC-insensitive mask’). Abbreviations: PCA=principal component analysis, NT=neurotypical, ASC=autism, F=females, M=males, RAP=region-aligned prediction method.

#### **SF7 – Comparison of highly mis- and accurately classified autistic individuals**

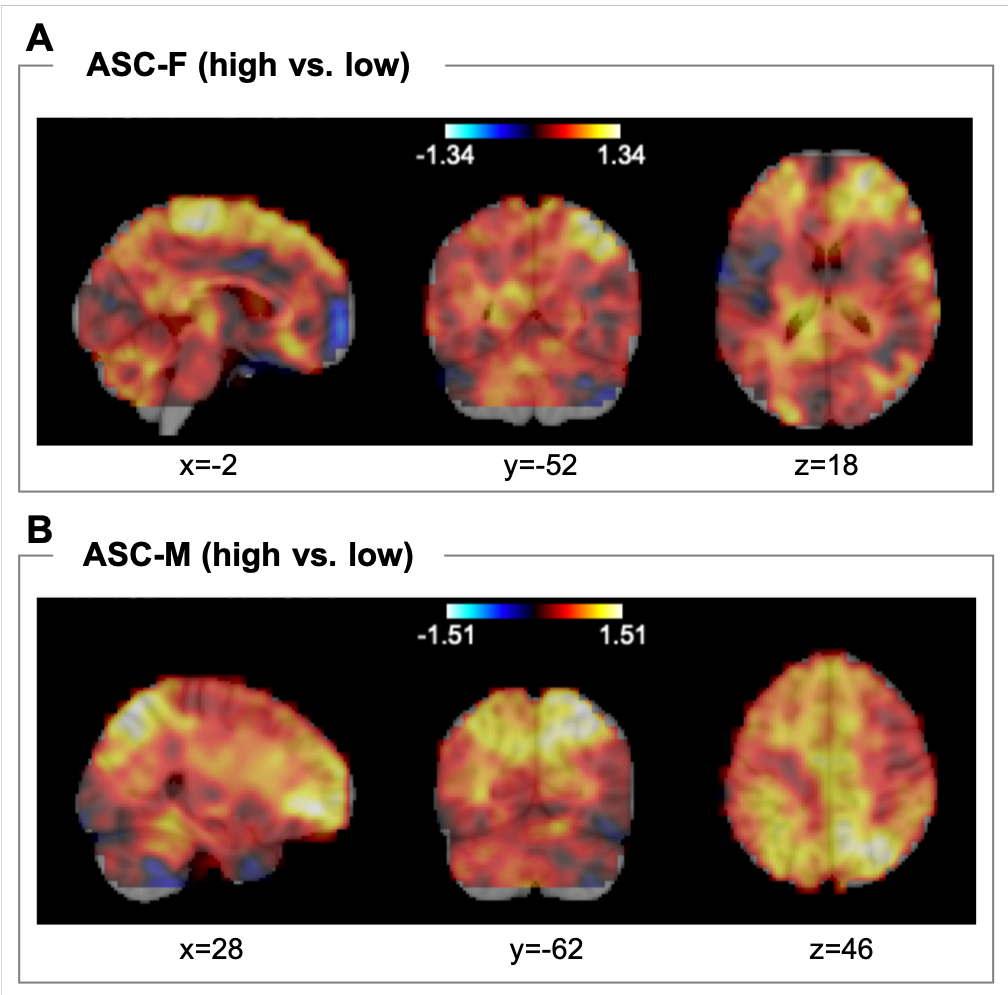

**SF7**. The brain maps depict the different RAP (spatial representation of the sex predictions) imaging-t-maps across **A**) highly-accurately classified vs. highly mis-classified autistic females and **B**) highly-accurately classified vs. highly mis-classified autistic males. Abbreviations: ASC=autism, F=females, M=males.

#### **SF8 – Enrichment analyses with more conservative threshold**

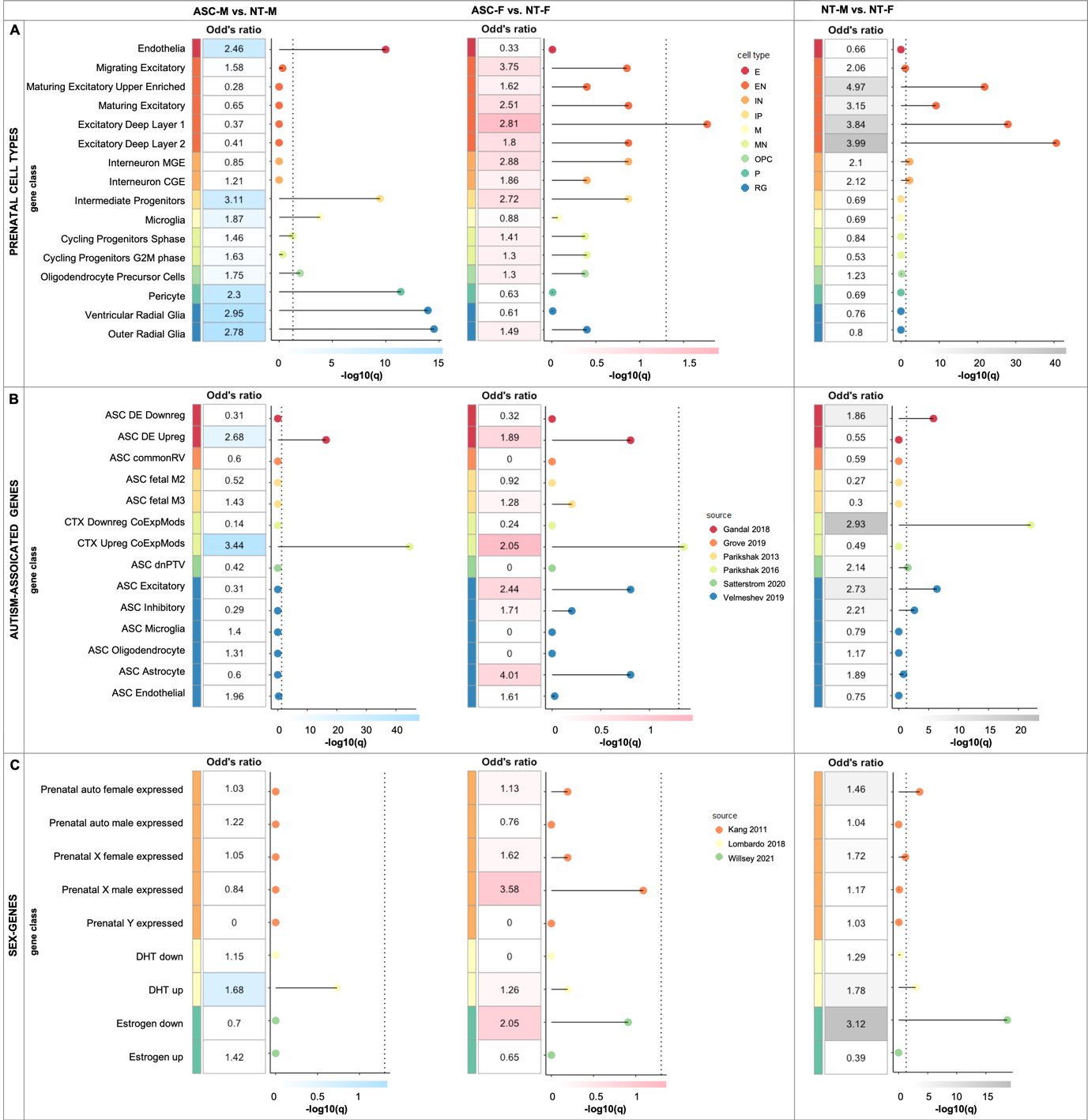

**SF8.** Enrichment analysis using a more conservative list of 16,906 background genes based on real estimates of genes expressed in cortical tissue(70). Odds-ratios at an FDR-corrected *P*<0.05 resulting from the gene set enrichment analyses of RAP-imaging-t-maps (ASC-M vs. NT-M; ASC-F vs. NT-F; NT-M vs. NT-F) and associated gene lists with different classes of genes acting prenatally and relevant in the context of autism and sexual differentiation. (**A**) genes from parental cell types(56) (endothelia (E), excitatory neurons (EN; migrating excitatory, maturing excitatory, maturing excitatory upper enriched, excitatory deep layer 1, excitatory deep layer 2), interneurons (IN; interneuron MGE and interneuron CGE), intermediate progenitors (IP), microglia (M), cycling progenitors (MN; cycling progenitor S-phase and cycling progenitors G2M phase), oligodendrocyte precursors (OPC), pericyte (P), and radial glia (RG; ventricular radial glia and outer radial glia), (**B**) autism-associated genes, including common genetic variants (ASC commonRV)(57), de novo mutations (fetal gene co-expression modules [ASC fetal M2, ASC fetal M3](60); 102 rare, de novo protein truncating genes [ASC dnPTV])(59) and transcriptionally dysregulated genes (differentially-expressed downregulated [ASC DE Downreg], differentially-expressed upregulated [ASD DE Upreg](61); cortical downregulated co-expression modules [ASC CTX Downreg CoExpMods], cortical upregulated co-expression modules [ASC CTX Upreg CoExpMods](62); ASC Excitatory, ASC Inhibitory, ASC Microglia, ASC Oligodendrocyte, ASC Astrocyte, ASC Endothelial(63)), and (**C**) sex-differentially expressed genes acting prenatally such as differentially regulated genes by dihydrotestosterone (DHT)(66–68), estrogen(69) and (autosomal and X-/Y-chromosome-linked) sex-differential gene array expression data from prenatal samples(64, 65). Abbreviations: NT=neurotypical, ASC=autism, F=females, M=males.

| **Variable** | | **ASC mean (std) [min-max]** | | **ASC mean (std) [min-max]** | | **NT M mean (std) [min-max]** | | **NT F mean (std) [min-max]** | | **post hoc** |
| --- | --- | --- | --- | --- | --- | --- | --- | --- | --- | --- |
| **N** | | 526 | | 115 | | 532 | | 239 | | - |
| **age** | | 13.2(5.7) [5.32-55.0] | | 13.66(7.28) [5.22-54.0] | | 13.53(6.27) [5.89-56.0] | | 13.07(6.36) [5.91-47.0] | | ASC=NT |
| **FIQ** | | 105.39(17.63) [41.0-149.0] | | 104.35(16.67) [66.0-146.0] | | 112.39(12.65) [71.0-148.0] | | 113.67(13.12) [80.0-149.0] | | ASC<NT |
| **VIQ** | | 106.3(18.59) [42.0-180.0] | | 105.71(16.67) [70.0-145.0] | | 113.71(13.47) [67.0-147.0] | | 114.09(14.56) [83.0-156.0] | | ASC<NT |
| **PIQ** | | 104.74(17.33) [37.0-149.0] | | 102.21(18.11) [53.0-148.0] | | 108.78(13.92) [62.0-147.0] | | 109.49(13.26) [79.0-145.0] | | ASC<NT |
| **ADI social** | | 19.64(5.34) [0.0-30.0] | | 19.08(6.2) [0.0-30.0] | | - | | - | | M=F |
| **ADI communication** | | 15.73(4.48) [0.0-25.0] | | 14.9(5.05) [0.0-24.0] | | - | | - | | M=F |
| **ADI RRB** | | 5.93(2.49) [0.0-12.0] | | 5.78(2.55) [0.0-12.0] | | - | | - | | M=F |
| **ADOS-G total** | | 11.38(3.97) [2.0-22.0] | | 11.81(3.9) [3.0-21.0] | | - | | - | | M=F |
| **ADOS-G coommunnication** | | 3.52(1.57) [0.0-8.0] | | 3.5(1.59) [0.0-7.0] | | - | | - | | M=F |
| **ADOS-G social** | | 7.74(2.78) [2.0-14.0] | | 7.95(2.45) [2.0-14.0] | | - | | - | | M=F |
| **ADOS-G RRB** | | 2.11(1.54) [0.0-7.0] | | 2.02(1.55) [0.0-5.0] | | - | | - | | M=F |
| **ADOS-2 CSS** | | 7.04(2.06) [1.0-10.0] | | 6.69(1.72) [2.0-10.0] | | - | | - | | M=F |
| **ADOS-2 SA** | | 9.37(3.81) [1.0-20.0] | | 8.79(3.14) [4.0-18.0] | | - | | - | | M=F |
| **ADOS-2 RRB CSS** | | 3.11(1.82) [0.0-8.0] | | 2.79(1.46) [0.0-6.0] | | - | | - | | M=F |

### Supplemental Tables

#### **ST1 – ABIDE sample characterization**

Abbreviations: ASC=autism, NT=neurotypical, M=male, F=females, FIQ=full-scale IQ, PIQ=performance IQ, VIQ=verbal IQ, ADI=Autism Diagnostic Interview, ADOS=Autism Diagnostic Observtion

Schedule, CSS=calibrated severity score, SA=social-affect, RRB=restricted, repetitive behavior

#### **ST2 – ADHD sample characterization**

| **Variable** | **ADHD M mean(std) [min-max]** | **ADHD F mean(std) [min-max]** | **NT M mean(std) [min-max]** | **NT F mean(std) [min-max]** | **post hoc** |
| --- | --- | --- | --- | --- | --- |
| **N** | 324 | 130 | 225 | 208 | - |
| **age** | 13.27(3.4) [7.25-21.0] | 13.02(3.95) [7.35-26.0] | 12.55(2.82) [7.29-22.0] | 12.83(3.86) [7.33-24.0] | ADHD=NT |
| **FIQ** | 102.81(16.77) [55.0-149.0] | 100.66(16.06) [48.0-134.0] | 114.08(15.35) [58.0-158.0] | 112.54(14.15) [75.0-144.0] | ADHD<NT |
| **VIQ** | 105.09(14.78) [54.0-141.0] | 104.25(16.22) [65.0-138.0] | 113.06(13.59) [71.0-141.0] | 111.49(14.27) [71.0-146.0] | ADHD<NT |
| **PIQ** | 101.82(14.76) [70.0-143.0] | 103.86(13.65) [79.0-135.0] | 114.41(13.69) [83.0-138.0] | 107.71(13.69) [67.0-135.0] | ADHD<NT |

Abbreviations: ADHD=attention-deficti/hyper-activity disorder, NT=neurotypical, M=male, F=females, FIQ=full-scale IQ, PIQ=performance IQ, VIQ=verbal IQ

#### **ST3 – Summary of acquisition parameters across sites in EU-AIMS LEAP**

| **Site** | **Manufacturer** | **Model** | **Software Version** | **Acquisition sequence** | **Coverage** | **Slices** | **Thickness [mm]** | **Resolution [mm^3^]** | **TR [s]** | **TE [ms]** | **FA [°]** | **FOV** |
| --- | --- | --- | --- | --- | --- | --- | --- | --- | --- | --- | --- | --- |
| Cambridge | Siemens | Verio | Syngo MR B17 | Tfl3d1_ns | 256*256 | 176 | 1.2 | 1.1*1.1*1.2 | 2.3 | 2.95 | 9 | 270 |
| London | GE Medical systems | Discovery mr750 | LX MR DV23.1_V02_1317.c | SAG ADNI GO ACC SPGR | 256*256 | 196 | 1.2 | 1.1*1.1*1.2 | 7.31 | 3.02 | 11 | 270 |
| Mannheim | Siemens | TimTrio | Syngo MR B17 | MPRAGE ADNI | 256*256 | 176 | 1.2 | 1.1*1.1*1.2 | 2.3 | 2.93 | 9 | 270 |
| Nijmegen | Siemens | Skyra | Syngo MRD13 | Tfl3d1_16ns | 256*256 | 176 | 1.2 | 1.1*1.1*1.2 | 2.3 | 2.93 | 9 | 270 |
| Rome | GE Medical systems | Signa HDxt | 24/LX/MR HD16.0_V02_1131.a | SAG ADNI GO ACC SPGR | 256*256 | 172 | 1.2 | 1.1*1.1*1.2 | 5.96 | 1.76 | 11 | 270 |
| Utrecht | Philips Medical Systems | Achieva/ | 3.2.3, 3.2.3.1 | ADNI GO 2 | 256*256 | 170 | 1.2 | 1.1*1.1*1.2 | 6.76 | 3.1 | 9 | 270 |
|  |  | Ingenia CX |  |  |  |  |  |  |  |  |  |  |

#### **ST4 – Summary of sample sized employed at different stages of the model application**

| **Sample** | **Dataset** | **NT-F** | **NT-M** | **ASC-F** | **ASC-M** | **Total** |
| --- | --- | --- | --- | --- | --- | --- |
| **Train** | ABIDE | 141 | 141 | 0 | 0 | 282 |
| **Test** | ABIDE | 84 | 84 | 84 | 84 | 336 |
| **Test** | LEAP | 98 | 98 | 98 | 98 | 392 |
| **Test (auto age-matching)** | LEAP | 85 | 85 | 85 | 85 | 340 |
| **Sample** | **Dataset** | **NT-F** | **NT-M** | **ADHD-F** | **ADHD-M** | **Total** |
| **Test (manual age-matching)** | ADHD | 130 | 130 | 130 | 130 | 520 |
| **Test (auto age-matching)** | ADHD | 125 | 125 | 125 | 125 | 500 |

Abbreviations: ASC=autism, NT=neurotypical, M=male, F=females, ADHD=attention-deficti/hyper-activity disorder

#### **ST5 – Sex-specific prediction accuracies in ADHD**

| **Cohort** | | **ADHD200 + NeuroImage** | | |
| --- | --- | --- | --- | --- |
| **Group** | | NT | ADHD-F | ADHD-M |
| **A – Manual age-matching *before* the model application** | | | | |
| **Sex-specific**  **accuracy** | Mean | 0.733 | 0.722 | 0.73 |
|  | SD | 0.046 | 0.069 | 0.064 |
| **B – Auto-age-matching *within* the model application** | | | | |
| **Sex-specific**  **accuracy** | Mean | 0.73 | 0.721 | 0.711 |
|  | SD | 0.045 | 0.066 | 0.062 |

Abbreviations: ADHD=attention-deficti/hyper-activity disorder, NT=neurotypical, M=male, F=females

#### **ST6 – Comparison of autistic individuals with and without ADHD**

| **Group** | | **NT** | **ASC-F** | | **ASC-M** | |
| --- | --- | --- | --- | --- | --- | --- |
| **Sub-group** | | **-** | **ADHD+** | **ADHD-** | **ADHD+** | **ADHD-** |
| **Sex-specific**  **accuracy** | Mean | 0.798718 | 0.714 | 0.747 | 0.866 | 0.863 |
|  | SD | 0.045 | 0.11 | 0.109 | 0.092 | 0.086 |

Abbreviations: ASC=autism, NT=neurotypical, M=male, F=females, ADHD=attention-deficti/hyper-activity disorder, ADHD+= autistic individuals with ADHD, ADHD-=autistic indivduals without ADHD

#### **ST7 – Sex-specific prediction accuracies across control analyses**

| **Group** | **NT** | | **ASC-F** | **ASC-M** |
| --- | --- | --- | --- | --- |
| **A – Auto-age-matching** | | | | |
| **Sex-specific**  **accuracy** | Mean | 0.8 | 0.737041 | 0.863265 |
|  | SD | 0.024182 | 0.052989 | 0.040996 |
| **B – Auto-age-matching + brain volume** | | | | |
| **Sex-specific**  **accuracy** | Mean | 0.733636 | 0.655455 | 0.843182 |
|  | SD | 0.05052 | 0.100125 | 0.06243 |
| **C – Principal component analysis and logistic regression** | | | | |
| **Sex-specific**  **accuracy** | Mean | 0.647 | 0.568 | 0.686 |
|  | SD | 0.037 | 0.049 | 0.052 |
| **D – RAP verification** | | | | |
| **Original** | Mean | 0.785 | 0.725238 | 0.814524 |
| **sex-specific**  **accuracy** | SD | 0.028193 | 0.047243 | 0.047136 |
| **ASC-sensitive mask** | mean | 0.701429 | 0.620476 | 0.762619 |
| **sex-specific**  **accuracy** | SD | 0.029539 | 0.055508 | 0.056015 |
| **ASC-insensitive mask** | Mean | 0.659762 | 0.638095 | 0.701667 |
| **sex-specific**  **accuracy** | SD | 0.036562 | 0.060272 | 0.062645 |

Abbreviations: ASC=autism, NT=neurotypical, M=male, F=females, RAP=region-aligned prediction

#### **ST8 – Most differentiating regions between NT M and NT F**

| **Grey Matter region** | **hemisphere** | **size** | **mean** | **std** |
| --- | --- | --- | --- | --- |
| Temporal Fusiform Cortex, posterior division | B | 22748 | 0.674848 | 0.155339 |
| Planum Temporale | L | 12209 | 0.667778 | 0.121297 |
| Caudate | L | 3949 | 0.602529 | 0.136994 |
| Heschl's Gyrus (includes H1 and H2) | L | 6084 | 0.676093 | 0.13534 |
| Parietal Operculum Cortex | B | 12782 | 0.661188 | 0.118787 |
| Parahippocampal Gyrus, posterior division | B | 14834 | 0.594444 | 0.130176 |
| Hippocampus | R | 6165 | 0.595922 | 0.090629 |
| Supramarginal Gyrus, posterior division | B | 32114 | 0.611748 | 0.10221 |
| Cerebellum Crus I | L | 26823 | 0.586231 | 0.108581 |
| Cerebellum Vermis Crus I | B | 5 | 0.59817 | 0.006328 |
| Cerebellum Vermis VI | B | 3736 | 0.581022 | 0.055262 |
| Temporal Occipital Fusiform Cortex | B | 19047 | 0.58575 | 0.125005 |
| Superior Temporal Gyrus, posterior division | B | 17277 | 0.610472 | 0.100785 |
| Angular Gyrus | B | 29354 | 0.580459 | 0.095334 |
| Planum Polare | B | 9013 | 0.565302 | 0.094504 |
| Inferior Temporal Gyrus, posterior division | B | 32133 | 0.559189 | 0.118362 |
| Accumbens | L | 756 | 0.551552 | 0.067611 |
| Supramarginal Gyrus, anterior division | B | 22159 | 0.590058 | 0.130507 |
| Cerebellum I-IV | R | 9228 | 0.558872 | 0.109159 |
| Central Opercular Cortex | B | 19707 | 0.530095 | 0.160716 |
| **White Matter connection** | **hemisphere** | **size** | **mean** | **std** |
| Sagittal stratum (include inferior longitidinal fasciculus and inferior fronto-occipital fasciculus) | R | 2228 | 0.744193 | 0.07272 |
| Sagittal stratum (include inferior longitidinal fasciculus and inferior fronto-occipital fasciculus) | L | 2231 | 0.694 | 0.072734 |
| Tapetum | L | 600 | 0.666728 | 0.032677 |
| Retrolenticular part of internal capsule | R | 2515 | 0.636811 | 0.095918 |
| Medial lemniscus | R | 690 | 0.662763 | 0.049701 |
| Tapetum | R | 596 | 0.636833 | 0.04043 |
| Cingulum (hippocampus) | R | 1236 | 0.60971 | 0.1172 |
| Retrolenticular part of internal capsule | L | 2469 | 0.569462 | 0.114106 |
| Posterior corona radiata | R | 3728 | 0.591619 | 0.065996 |
| Corticospinal tract | R | 1362 | 0.580639 | 0.067095 |
| Fornix (cres) / Stria terminalis (can not be resolved with current resolution) | R | 1124 | 0.574191 | 0.062119 |
| Anterior limb of internal capsule | L | 3018 | 0.558917 | 0.085043 |
| Cingulum (hippocampus) | L | 1155 | 0.564226 | 0.071553 |
| Genu of corpus callosum | B | 8851 | 0.542123 | 0.104349 |
| Posterior corona radiata | L | 3714 | 0.547375 | 0.072429 |
| Splenium of corpus callosum | B | 12729 | 0.539905 | 0.056821 |
| Cerebral peduncle | R | 2278 | 0.530177 | 0.063799 |
| Inferior cerebellar peduncle | R | 968 | 0.536343 | 0.060001 |
| Posterior thalamic radiation (include optic radiation) | L | 3978 | 0.515662 | 0.16168 |

Abbreviations: NT=neurotypical, M=male, F=female, R=right, L=left, B=both

#### **ST9 – Most male-shifted regions in ASC F**

| **Grey Matter region** | **hemisphere** | **size** | **mean** | **std** |
| --- | --- | --- | --- | --- |
| Cerebellum Crus I | L | 26823 | 0.255442 | 0.072154 |
| Heschl's Gyrus (includes H1 and H2) | B | 6084 | 0.226662 | 0.104109 |
| Caudate | L | 3949 | 0.188232 | 0.110903 |
| Planum Temporale | L | 12209 | 0.183346 | 0.134065 |
| Occipital Fusiform Gyrus | B | 28100 | 0.204175 | 0.133693 |
| Amygdala | L | 2662 | 0.168623 | 0.063565 |
| Parietal Operculum Cortex | B | 12782 | 0.171952 | 0.086195 |
| Hippocampus | L | 6154 | 0.165365 | 0.0567 |
| Accumbens | L | 756 | 0.171228 | 0.040709 |
| Inferior Temporal Gyrus, temporooccipital part | B | 20013 | 0.157296 | 0.083235 |
| Cerebellum VI | L | 17861 | 0.172028 | 0.094903 |
| Inferior Temporal Gyrus, posterior division | B | 32133 | 0.146074 | 0.114158 |
| Caudate | R | 4127 | 0.142831 | 0.092932 |
| Cerebellum Crus II | L | 21227 | 0.141174 | 0.089322 |
| Central Opercular Cortex | B | 19707 | 0.132469 | 0.113722 |
| Temporal Occipital Fusiform Cortex | B | 19047 | 0.137184 | 0.08119 |
| Lateral Occipital Cortex, inferior division | B | 57896 | 0.139874 | 0.107609 |
| Superior Temporal Gyrus, posterior division | B | 17277 | 0.122037 | 0.156537 |
| Planum Polare | B | 9013 | 0.132844 | 0.079657 |
| Parahippocampal Gyrus, posterior division | B | 14834 | 0.116739 | 0.086311 |
| **White Matter connection** | **hemisphere** | **size** | **mean** | **std** |
| Genu of corpus callosum | B | 8851 | 0.272063 | 0.102349 |
| Retrolenticular part of internal capsule | R | 2515 | 0.27698 | 0.06932 |
| Sagittal stratum (include inferior longitidinal fasciculus and inferior fronto-occipital fasciculus) | R | 2228 | 0.221877 | 0.115045 |
| Tapetum | L | 600 | 0.203539 | 0.110326 |
| Retrolenticular part of internal capsule | L | 2469 | 0.218705 | 0.063159 |
| Fornix (cres) / Stria terminalis (can not be resolved with current resolution) | L | 1125 | 0.207553 | 0.044443 |
| Sagittal stratum (include inferior longitidinal fasciculus and inferior fronto-occipital fasciculus) | L | 2231 | 0.195957 | 0.039052 |
| Tapetum | R | 596 | 0.199917 | 0.020729 |
| Fornix (cres) / Stria terminalis (can not be resolved with current resolution) | R | 1124 | 0.17257 | 0.101445 |
| Uncinate fasciculus | L | 376 | 0.164049 | 0.029604 |
| Superior longitudinal fasciculus | R | 6607 | 0.167562 | 0.059507 |
| Superior cerebellar peduncle | R | 992 | 0.137128 | 0.092553 |
| Posterior thalamic radiation (include optic radiation) | L | 3978 | 0.119328 | 0.145831 |
| Cingulum (hippocampus) | L | 1155 | 0.130622 | 0.023392 |
| Posterior corona radiata | R | 3728 | 0.13005 | 0.034196 |
| Posterior thalamic radiation (include optic radiation) | R | 3972 | 0.138363 | 0.095073 |
| Uncinate fasciculus | R | 380 | 0.120574 | 0.022961 |
| Medial lemniscus | R | 690 | 0.137374 | 0.06416 |
| Corticospinal tract | R | 1362 | 0.132608 | 0.080538 |

Abbreviations: ASC=autism, F=female, R=right, L=left, B=both

#### **ST10 – Most male-shifted regions in ASC M**

| **Grey Matter region** | **hemisphere** | **size** | **mean** | **std** |
| --- | --- | --- | --- | --- |
| Amygdala | R | 3215 | 0.17297 | 0.054697 |
| Pallidum | R | 2118 | 0.164403 | 0.02943 |
| Accumbens | R | 666 | 0.159551 | 0.022322 |
| Thalamus | L | 11488 | 0.158337 | 0.033188 |
| Cerebellum VIIIa | R | 9826 | 0.137376 | 0.070234 |
| Cingulate Gyrus, anterior division | B | 31689 | 0.149518 | 0.04923 |
| Thalamus | R | 11186 | 0.14557 | 0.035606 |
| Putamen | R | 6397 | 0.132497 | 0.046621 |
| Caudate | R | 4127 | 0.130855 | 0.027515 |
| Subcallosal Cortex | B | 16466 | 0.131314 | 0.047305 |
| Cerebellum VIIb | R | 9885 | 0.115508 | 0.065721 |
| Frontal Medial Cortex | B | 12100 | 0.112058 | 0.070862 |
| Cerebellum VIIIb | R | 8185 | 0.114916 | 0.055648 |
| Juxtapositional Lobule Cortex (formerly Supplementary Motor Cortex) | B | 17545 | 0.113293 | 0.042752 |
| Pallidum | L | 2133 | 0.105007 | 0.049254 |
| Cingulate Gyrus, posterior division | B | 35642 | 0.100388 | 0.051582 |
| Temporal Pole | B | 62256 | 0.095262 | 0.061998 |
| Accumbens | L | 756 | 0.097188 | 0.027359 |
| Amygdala | L | 2662 | 0.0988 | 0.055703 |
| Paracingulate Gyrus | B | 31069 | 0.096296 | 0.052689 |
| **White Matter connection** | **hemisphere** | **size** | **mean** | **std** |
| Superior corona radiata | R | 7500 | 0.195155 | 0.033281 |
| Superior fronto-occipital fasciculus (could be a part of anterior internal capsule) | R | 507 | 0.191542 | 0.009216 |
| Anterior corona radiata | R | 6849 | 0.172089 | 0.041197 |
| Cerebral peduncle | L | 2278 | 0.166158 | 0.034835 |
| Cingulum (cingulate gyrus) | R | 2342 | 0.168892 | 0.041982 |
| Posterior limb of internal capsule | R | 3754 | 0.154056 | 0.041451 |
| Genu of corpus callosum | B | 8851 | 0.143966 | 0.062544 |
| Cerebral peduncle | R | 2278 | 0.140164 | 0.082683 |
| Body of corpus callosum | B | 13711 | 0.143612 | 0.033902 |
| Posterior limb of internal capsule | L | 3752 | 0.137433 | 0.026209 |
| Superior longitudinal fasciculus | R | 6607 | 0.128455 | 0.082359 |
| Anterior limb of internal capsule | R | 3138 | 0.14209 | 0.02755 |
| Fornix (column and body of fornix) | B | 659 | 0.128445 | 0.031074 |
| Retrolenticular part of internal capsule | L | 2469 | 0.122443 | 0.055491 |
| Fornix (cres) / Stria terminalis (can not be resolved with current resolution) | L | 1125 | 0.121074 | 0.058493 |
| Superior fronto-occipital fasciculus (could be a part of anterior internal capsule) | L | 507 | 0.121725 | 0.033861 |
| Superior corona radiata | L | 7508 | 0.111048 | 0.041673 |
| Cingulum (cingulate gyrus) | L | 2751 | 0.112687 | 0.035577 |

Abbreviations: ASC=autism, M=male, R=right, L=left, B=both
